# Early-life exposure to the Chinese famine of 1959-61 and type 2 diabetes in adulthood: a systematic review and meta-analysis

**DOI:** 10.1101/2022.02.16.22271081

**Authors:** Chihua Li, L.H. Lumey

## Abstract

**Background:** The Chinese famine of 1959–61 has been widely interpreted as an important driver of current and future type 2 diabetes (T2D) epidemics. We conducted a systematic review and meta-analysis of prenatal famine exposure and type 2 diabetes (T2D) in China to summarize study characteristics, examine impacts of control selections and other selected characteristics on study results, identify other characteristics influencing results, and formulate recommendations for future studies.

**Methods:** We searched English and Chinese databases for studies that examined the relationship between T2D and prenatal exposure to the Chinese famine up to February 8^th^, 2022. From included studies, we extracted information on the number of T2D cases and populations at risk among individuals born during the famine (famine births), before the famine (pre-famine births), and after the famine (post-famine births). We compared risk of T2D in famine births to different controls: post-famine births, pre- and post-famine births combined, and pre-famine births. Heterogeneity across studies was assessed, and random-effects models were used to calculate summary estimates. Meta-regressions were used to examine the relationship between effect estimates and age differences. Subgroup analyses were performed based on selected characteristics, including participants’ sex, age, T2D measurement, famine intensity, residence, and publication language.

**Findings:** In total, 23 studies met our inclusion criteria. Sample sizes ranged from below 300 to over 350,000. All studies defined famine exposure based on participants’ date of birth, and 18 studies compared famine births to controls of post-famine births to estimate famine effects on T2D. Famine and post-famine births had an age difference of three years and over in each study. Using post-famine births as controls, a random-effects model shows an increased risk of T2D (OR 1.50, 95% CI 1.34–1.68) among famine births. In contrast, a marginally increased risk of T2D (OR 1.12, 95% CI 1.02–1.24) can be observed using pre- and post-famine births combined as controls, and a decreased risk (OR 0.89, 95% CI 0.79–1.00) using pre-famine births as controls. Studies with larger age differences between comparison groups had larger famine effects. Effect estimates comparing famine births to pre- and post-famine births combined depend on none of above selected characteristics. Studies showed a large variation in sampling sources, famine intensity assessment, and confounding adjustment.

**Interpretation:** Current estimates of a positive relation between prenatal exposure to the Chinese famine and adult T2D are mainly driven by uncontrolled age differences between famine births and post-famine controls. Marginal or no effects remain after controlling for the differences in most Chinese famine studies. It remains an open question to what extent the famine is related to current T2D patterns in China. Studies with more rigorous methods including age-balanced controls and robust famine intensity measures will be needed to quantify this relationship.

**Funding:** None.

## INTRODUCTION

Famines in human history provide unique opportunities to study how early-life environments may influence health outcomes.^1,2^ In the past two decades, scholars became increasingly interested in assessing the long-term impacts of early-life exposure to the Great Chinese Famine of 1959-61 (Chinese famine) on health outcomes.^3-5^ Five years ago, we conducted a systematic review and meta-analysis of 36 Chinese famine studies to summarize the data and generate estimates of homogeneity of reported effects.^4^ It was the first meta-analysis on famine exposure in China and health outcomes, including overweight/obesity, diabetes/hyperglycemia, hypertension, metabolic syndrome and schizophrenia. It included eight studies that reported findings about diabetes and/or glucose dysregulation. We found that Chinese famine studies predominantly compared individuals born during the famine to controls born after the famine. Uncontrolled age differences between comparison groups can explain ‘apparent’ famine effects because the risk of most chronic conditions increased with age.

After our review, many new famine studies have been conducted. By 2021, there are almost 200 original studies relating early-life Chinese famine exposure to adverse health outcomes in adulthood (**Figure 1**), including overweight/obesity, hypertension, type 2 diabetes (T2D), metabolic syndrome, cardiovascular disease, cancer, psychological disorder, and many others. Among them, T2D was the most widely examined outcome, with a number of original studies over 30. In addition, multiple meta-analyses have been conducted to summarize long-term impacts of the Chinese famine and other famines on health outcomes.^6-12^ These individual studies and meta-analyses claimed increased risks of adverse health outcomes with prenatal famine exposure. Many reviews and commentaries therefore concluded that famine exposure is a major driver for the current T2D epidemic in China and will contribute to an increased risk of T2D in future generations.^3,13-20^

**Figure 1.**
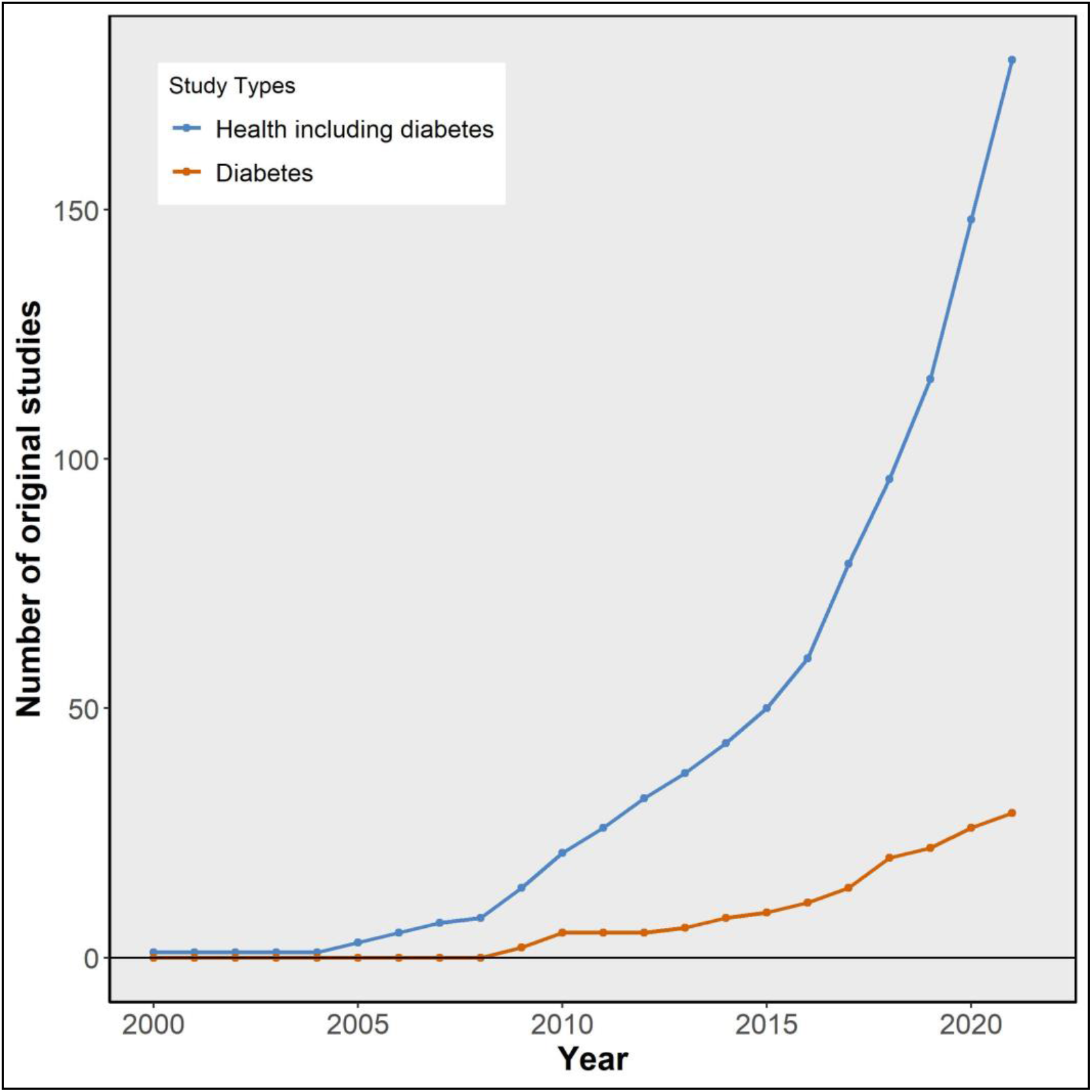
Cumulative number of original research articles on the Chinese famine and health outcomes by year. A full list of original research articles on the Chinese famine and health outcomes published in 2021 and before can be found in the Supplementary Reference List.

We have previously shown that most of the Chinese famine studies conducted before 2017 had major methodological problems, including uncontrolled age difference, poor famine intensity assessment, and biased convenience sampling.^4^ It is unclear how later Chinese famine studies have been conducted and if improvements are made. Considering the topic’s increasing importance, we again conducted a systematic review and meta-analysis of Chinese famine studies on T2D to summarize study characteristics and update results. We used the meta-analysis as a comparative tool to examine the impact of control selections on study results and to identify other characteristics affecting study results.^21-23^ We quantified the relationship between age differences of comparison groups and reported famine effect across studies. We conducted a quality assessment and provided specific recommendations for future studies.

## METHODS

### Search strategy and study selection

We followed the Preferred Reporting Items for Systematic Reviews and Meta-Analyses (PRISMA) guidelines (**Supplementary Table 1**).^24^ The study protocol is presented in **Supplementary Text 1**. Five electronic databases in English and Chinese languages were searched for Chinese famine studies on T2D up to February 8^th^ 2022, including PubMed, Embase, Web of Science, Wanfang Data, and the Chinese National Knowledge Infrastructure (CNKI). Broad search terms in English and Chinese were used to capture related studies, including journal articles, degree theses, and conference manuscripts. The following keywords were searched: [((China OR Chinese) AND (famine OR undernutrition OR starvation OR malnutrition)) OR great leap forward OR great famine]. Review articles and reference lists were screened for relevant studies.

Studies meeting the following criteria were included: (a) the study was reported as original research work; (b) the Chinese famine of 1959-61 was the exposure of interest; (c) T2D, or hyperglycemia, or increased blood glucose was the outcome of interest; (d) information on study design and results was provided. The full texts of relevant studies were examined to determine if they met the inclusion criteria. When several studies were available on the same or overlapping cohorts reporting results of T2D and glucose dysregulation, we selected results from studies that either provided the most comprehensive information or had the largest sample size as representatives. Our search identified 14 Chinese famine studies on T2D using the same or overlapping data sources, including the Kailuan Group Health Examination,^25,26^ China Kadoorie Biobank (CKB),^27,28^ China National Nutrition and Health Survey (CNNHS) in 2010-12,^29,30^ Survey on Prevalence in East China for Metabolic Diseases and Risk Factors Cohort (SPECT) in Shanghai, Jiangxi and Zhejiang,^31-33^ China Health and Retirement Longitudinal Studies (CHARLS).^34-38^ From them, five studies were selected as representative studies in our systematic review and meta-analysis.^25,27,35^

### Data extraction and quality assessment

The following data were extracted from included studies: author and publication information, study characteristics, time windows used to define different comparison groups, and tabular information on the number of T2D cases and populations at risk (**Supplementary Text 1**). Among included studies, seven studies provided only famine effect estimates but no information on the number of T2D cases and populations at risk for famine births or post-famine births.^12,25,39-43^ Following a previous study, a modified Newcastle-Ottawa scale was used to evaluate three domains (sample, design, and analysis) containing eight items in total for each included study: sampling source, sample size, outcome assessment, exposure definition, control selection, famine intensity assessment, confounding adjustment, and statistical analysis (**Supplementary Text 2**).^4,44^ The quality of each item was scored ‘good (2)’, ‘fair (1)’ or ‘poor (0)’ based on predefined criteria for each study, and a total score was calculated (range: 0-16). Two reviewers (C.L. and L.H.L) appraised each study independently.

### Statistical analysis

For consistency, participants born during the Chinese famine of 1959-61 were defined as famine births (prenatally exposed); participants born after the famine were defined as post-famine births; and participants born before the famine were defined as pre-famine births. Whenever possible, pre-famine births and post-famine births were combined as a single control group. Age differences were further calculated by comparing famine births to different comparison groups, including post-famine births, pre- and post-famine births combined, and pre-famine births.

Packages of *meta* and *metafor* in R 4.1.0 were used to perform the meta-analysis.^45^ For each study, odds ratios (ORs) and 95% confidence intervals (CIs) for T2D were calculated by comparing famine births to different control groups: post-famine births (the most commonly used control group in Chinese famine studies), pre- and post-famine births combined, and pre-famine births. This will show how study results may change based on the selection of control groups. Fixed-effect (Mantel-Haenszel) model and random-effects (Dersimonian-Laird) models were used to obtain summary effect estimates (ORs and 95% CIs).^46^ The I^2^ statistic was used to estimate the percentage of variability across reports. To examine the influence of each study on meta-analysis results, leave-one-out analysis was conducted by omitting one study at the time and repeating the meta-analysis separately. To identify potential characteristics influencing study results, subgroup analyses were performed by sex, mean age at the survey, T2D measurements, reported famine intensity, urban/rural residence, and publication language.^23,47^ Publication bias was assessed by funnel plots and Egger’s regression test.^45,48^

## RESULTS

### Study characteristics

The above search strategy identified 47,709 records from database searches and other sources (**Figure 2**). After the removal of duplicates and title/abstract screening, 78 studies were selected for full-text review. This yielded 23 Chinese famine studies on T2D meeting the inclusion criteria.^25,27,30,31,35,39-43,49-61^ **Table 1** summarizes their selected characteristics: authors, language, data source, outcome assessment, control selection, and reported results. Fourteen of these studies were in English, and the rest were in Chinese. Eighteen studies compared famine births to post-famine births to estimate famine effects, and five studies compared famine births to pre- and post-famine births combined. Most studies used ADA or WHO definitions to measure T2D,^62-64^ and reported a 1.2 to 2-fold increase in the odds of T2D except for one study reporting a 5.7-fold increase (Study #17).^56^

**Figure 2.**
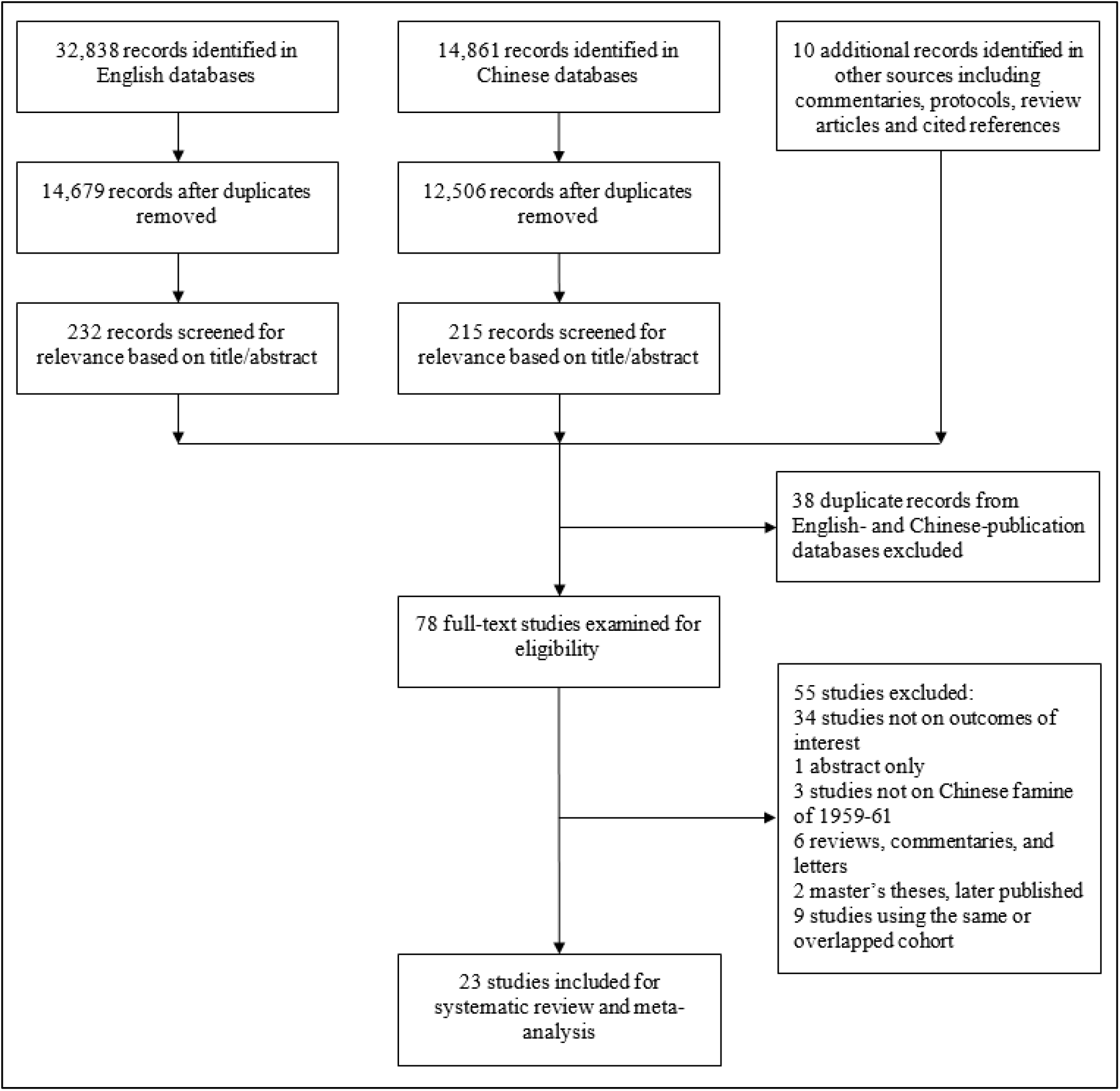
Flow diagram of study inclusion.

**Table 1.** Main characteristics of included Chinese famine studies on T2D.

| **Study #** | **Authors** | **Language** | **Data source** | **Outcome assessment** | **Control selection** | **Reported relationship of fetal famine exposure and T2D\*** |
| --- | --- | --- | --- | --- | --- | --- |
| 1 | Liu et al. 2009<sup>49</sup> | Chinese | Chongqing First Hospital Affiliated Health Examination Center, 2007 | Fasting blood glucose | Post | Increased level of fasting blood glucose and prevalence of T2DM |
| 2 | Guan et al. 2009<sup>39</sup> | Chinese | Chongqing Gangtie Group, 2009 | Fasting blood glucose | Post | Increased level of fasting blood glucose |
| 3 | Li et al. 2010<sup>50</sup> | English | China National Nutrition and Health Survey (CNNHS), 2002 | WHO 1998 | Post | OR: 1.43 (0.53, 3.87) for famine severe areas; 0.41 (0.12, 1.35) for famine less severe areas |
| 4 | Li et al. 2010<sup>41</sup> | Chinese | Chongqing First Hospital Affiliated Health Examination Center, 2010 | Fasting blood glucose | Pre and Post | Increased level of fasting blood glucose |
| 5 | Zhang et al. 2010<sup>40</sup> | Chinese | Tangshan Resident Study, 2009 | ADA 1997 | Pre and Post | OR: 1.69 (1.06, 2.69) |
| 6 | Zhao et al. 2013<sup>51</sup> | Chinese | Anhui Medical University Affiliated Health Examination Center, 2011 | WHO 1999 | Post | RR: 0.91 (0.37, 2.23) |
| 7 | Li et al. 2014<sup>25</sup> | Chinese | Kailuan Group, 2006-07 | WHO 1998 | Pre and Post | OR: 1.22 (1.06, 1.40) |
| 8 | Zhang et al. 2014<sup>42</sup> | Chinese | Bengbu First Hospital Affiliated Health Examination Center, 2011 | Fasting blood glucose | Post | No increased level of fasting blood glucose |
| 9 | Wang et al. 2015<sup>31</sup> | English | Survey on Prevalence in East China for Metabolic Diseases and Risk Factors Cohort (SPECT) in Shanghai, Jiangxi, Zhejiang, 2014 | ADA 2014 | Post | OR: 1.63 (1.13, 2.35) |
| 10 | Wang et al. 2016<sup>52</sup> | English | Dongfengtongji Cohort (DFTJ), 2008 | WHO 1998 and ADA 2010 | Post | OR: 1.03 (0.77, 1.38)<br>Same results using either WHO or ADA criteria |
| 11 | Wang et al. 2017<sup>53</sup> | English | Survey on Prevalence in East China for Metabolic Diseases and Risk Factors cohort (SPECT) in Anhui, 2014 | ADA 2014 | Post | OR: 1.90 (1.12, 3.21)<br>for famine severe areas |
| 12 | Li et al. 2017<sup>54</sup> | English | Suihua Cohort, 2015 | WHO 1999 | Post | OR: 1.75 (1.20, 2.54) |
| 13 | Meng et al. 2018<sup>27</sup> | English | China Kadoorie Biobank (CKB), 2004-8 | ICD-10: E12&14 | Post<sup>#</sup> | HR: 1.25 (1.07, 1.45) |
| 14 | Wang et al. 2018<sup>35</sup> | English | China Health and Retirement Longitudinal Study (CHARLS), 2011-12 | ADA 2017 | Pre and Post | OR: 1.37 (1.09, 1.72) |
| 15 | Zhang et al. 2018<sup>55</sup> | English | Chronic Disease Survey of Jilin Province, 2012 | WHO 1998 | Post<sup>#</sup> | OR: 1.51 (1.15, 1.98) |
| 16 | Zhou et al. 2018<sup>57</sup> | English | Hefei City Resident Study, 2011-12 | WHO 2006 | Post | RR: 0.72 (0.16, 3.33) |
| 17 | Liu et al. 2019<sup>56</sup> | Chinese | Guangxi Zhuang Nationality Resident Study, 2017 | ADA 2017 | Post | OR: 5.71 (1.53, 21.2) |
| 18 | Lu et al. 2020<sup>58</sup> | English | China Cardiometabolic Disease and Cancer Cohort (4C), 2011-16 | ADA 2017 | Post<sup>#</sup> | RR: 1.17 (1.05, 1.31) |
| 19 | Zhang et al. 2020<sup>30</sup> | English | China National Nutrition and Health Survey (CNNHS), 2010-12 | WHO 1999 | Post | OR: 1.31 (1.01, 1.70) |
| 20 | Qi et al. 2020<sup>43</sup> | Chinese | Shanghai Jiading Community, 2018 | WHO 1999 | Post | OR: 1.52 (1.07, 2.14)<br>for men; 1.74 (1.22, 2.50) for women |
| 21 | Ning et al. 2021<sup>59</sup> | English | Qingdao Diabetes Prevention program, 2006-09 | WHO 2006 | Post | RR: 2.15 (1.29, 3.60) |
| 22 | Zhang et al. 2022<sup>60</sup> | English | YiduCloud Clinic Data, 1999-2018 | Clinical records | Pre and Post | Increased prevalence of T2D among both males and females |
| 23 | Huo et al. 2022<sup>61</sup> | English | Henan Rural Cohort Study | WHO 1998 and ADA 2009 | Post | OR: 1.65 (1.29, 2.09) |

Pre: pre-famine births; Post: post-famine births
\* Association estimate based on fully adjusted model

Additional study information was summarized in **Supplementary Table 2**, including study design, sampling method, sample size, famine intensity measurement, analytical method, and covariate adjustment. Most studies analyzed data cross-sectionally except for two studies that followed participants’ T2D over time (Study #13 and 18).^27,58^ Eight surveys adopted hospital- or corporation-based convenience sampling, collecting data from patients or employees who had general health examinations in a single year.^25,39,41,42,49,51,52,60^ Other surveys or cohort studies used systematic sampling at both regional and national levels. The sample size varied from a few hundred to over 360,000. Seven studies measured famine intensity either based on mortality rates or grain production in the 1950-60s.^30,35,42,50,52-54^ Most studies used logistic regression to analyze the data. Twenty studies adjusted for different sets of covariates, and three studies did not make any adjustments.^41,42,49^

### Age differences and effect estimates comparing famine births to different controls

Mean age at the survey for different comparison groups are summarized in **Supplementary Table 3**, including pre-famine births, famine births, post-famine births, and pre- and post-famine births combined. Five studies did not provide any age information.^39,41-43,60^ Exact years and months of birth used to define famine births, pre-famine births, and post-famine births in each study are presented in **Supplementary Figure 1**. Four studies recruited famine and post-famine births but not pre-famine births.^30,39,54,56^ **Figure 3A** shows an age difference of three years or more comparing famine births to post-famine births or pre-famine births in each study. Age differences are reduced close to zero or no greater than one year comparing famine births to pre- and post-famine births combined in most studies except for three studies (Study #18, 21 and 23).

**Figure 3.**
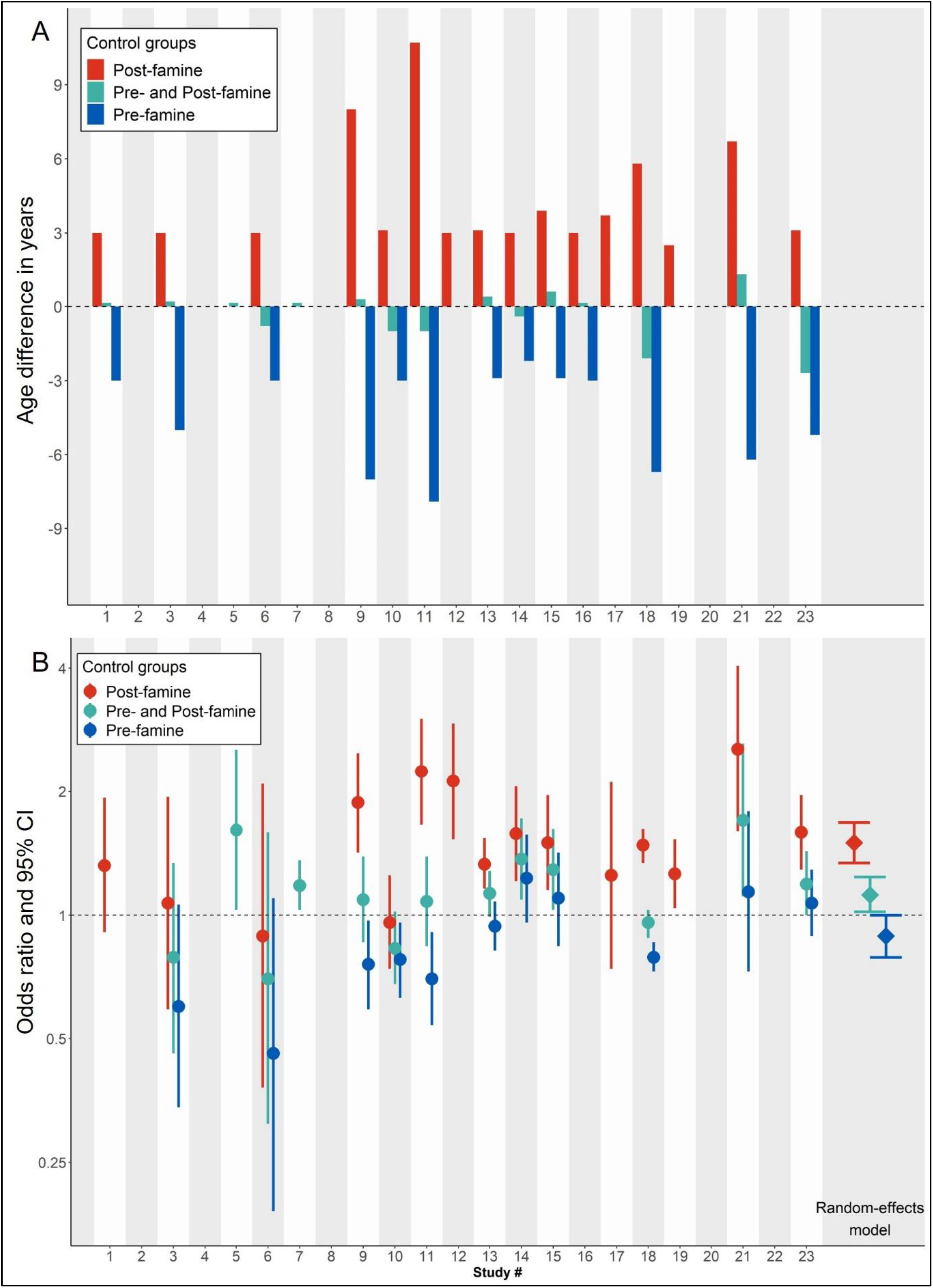
Age differences and effect estimates comparing famine births to different controls. A. Age differences comparing famine births to different control groups, including post-famine births, pre-famine births, and pre- and post-famine births combined. B. Effect estimates comparing famine births to different control groups. Odds ratio calculated based on number of T2D cases and population at risk for each individual study. Random-effects models used to calculate summary effect estimate for each control group.

### Meta-analysis comparing famine births to different control groups

Directions of famine effects on T2D are sensitive to selection of control groups. **Figure 3B** shows an increased odds of T2D (OR between 1.30 and 2.50) comparing famine births to post-famine births in most studies, in board agreement with reported negative famine effects in **Table 1**. The random-effects model summarizing individual estimates also shows a negative effect (OR 1.50, 95% CI 1.34-1.68). In contrast, comparing famine births to pre- and post-famine births combined shows a null effect or marginally increased odds of T2D in most studies and by the random-effects model (OR 1.12, 95% CI 1.02-1.24). Comparing famine births to pre-famine births even shows a ‘protective effect’ in most studies and by the random-effects model (OR 0.89, 95% CI 0.79-1.00). **Supplementary Figure 2A-C** shows detailed information of meta-analysis results comparing famine births to the three different control groups. Leave-one-out analysis shows that the meta-analysis results are robust by omitting one study at the time (**Supplementary Figure 3A-C**).

### Meta-regression and subgroup analysis

Magnitudes of famine effects on T2D increase with increasing age differences between comparison groups whichever control group is used (**Figure 4**). Comparing famine births to post-famine births, the famine effect increases by 1.07 times (95% CI 1.02-1.11) with one year increase in age difference; comparing famine births to pre- and post-famine births combined, the famine effect increases by 1.07 times (95% CI 0.98-1.07) with one year increase in age difference; comparing famine births to pre-famine births combined, the famine effect increases by 1.05 times (95% CI 1.00-1.11) with one year increase in age difference.

**Figure 4.**
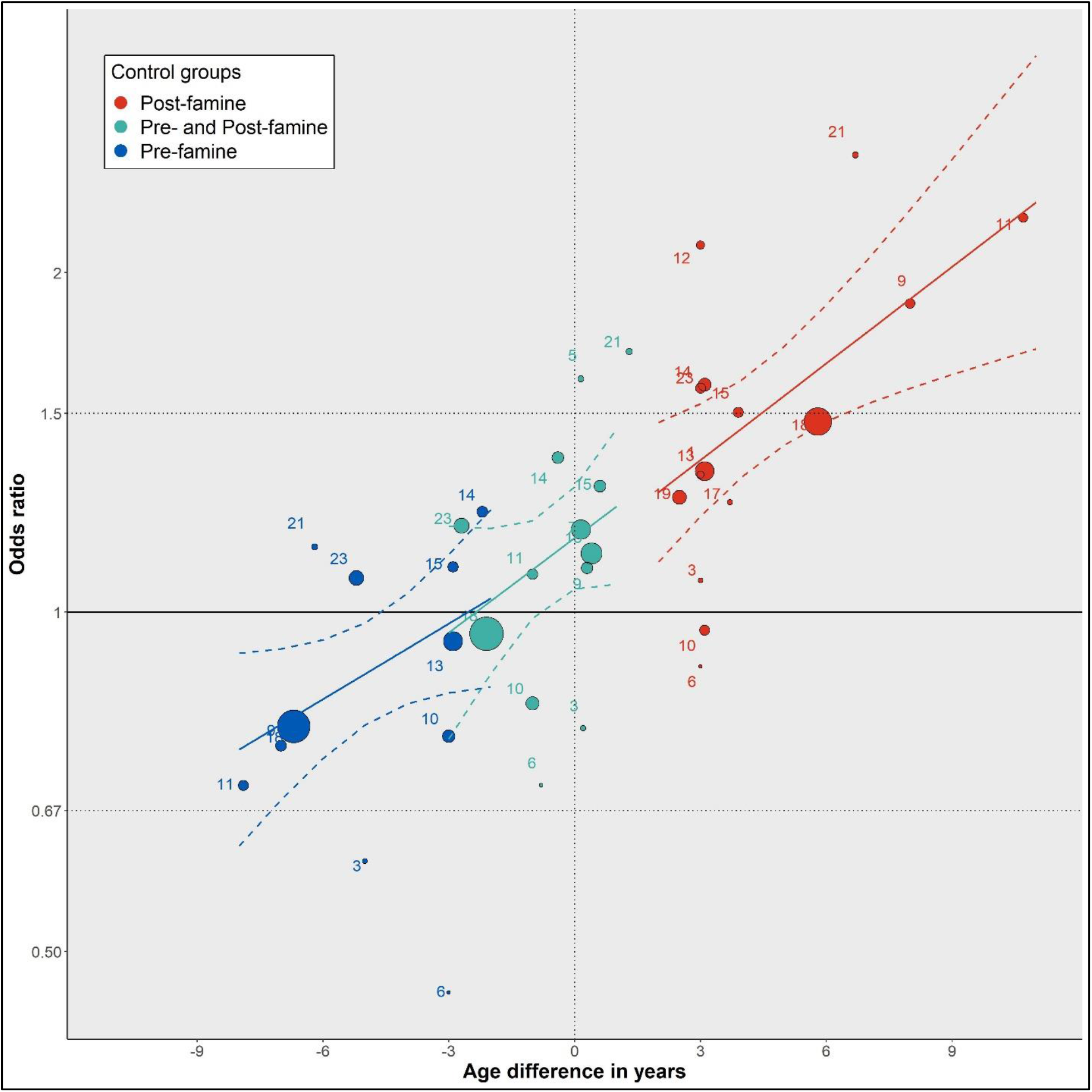
Meta-regressions of famine effect estimates over age differences comparing famine births to different control groups. The size of each dot is proportional to the weight of the study. The dashed lines in color represents 95%CI for each meta-regression model.

To identify potential characteristics affecting study results other than control selections and age differences, subgroup analysis of comparing famine births to pre- and post-famine births combined is conducted based on sex, mean age at survey, T2D measurements, reported famine intensity, residence, and publication language. **Table 2** shows that any famine effects on T2D are marginal after stratifications by these different characteristics (random-effects model OR between 0.95 and 1.25). Detailed results of stratified analysis are presented in **Supplementary Figure 4A-F**. Meta-regression using these selected characteristics shows consistent results from above subgroup analyses.

**Table 2.** Effect estimates of famine exposure on T2D comparing famine births with pre- and post-famine births combined after stratification by selected characteristics.

|  | **Fixed effect model** |  | **Random-effects model** |  |
| --- | --- | --- | --- | --- |
|  | OR | 95% CI | OR | 95% CI |
| **Sex** |  |  |  |  |
| Men | 1.22 | (1.11; 1.34) | 1.22 | (1.11; 1.34) |
| Women | 1.12 | (1.02; 1.23) | 1.22 | (1.02; 1.46) |
| Men and women mixed | 0.96 | (0.89; 1.03) | 0.96 | (0.89; 1.03) |
| **Mean age at survey** |  |  |  |  |
| <50 years | 1.21 | (1.08; 1.37) | 1.27 | (0.98; 1.66) |
| >=50 years | 1.04 | (0.98; 1.10) | 1.09 | (0.98; 1.21) |
| **T2D measurements** |  |  |  |  |
| WHO | 1.12 | (1.03; 1.22) | 1.11 | (0.94; 1.32) |
| ADA | 1.02 | (0.95; 1.09) | 1.14 | (0.96; 1.37) |
| ICD-10 | 1.13 | (0.99; 1.28) | 1.13 | (0.99; 1.28) |
| **Reported famine intensity** |  |  |  |  |
| Severe | 1.24 | (1.01; 1.53) | 1.25 | (1.00; 1.56) |
| Less severe | 1.18 | (1.07; 1.29) | 1.18 | (1.03; 1.34) |
| Mixed | 1.01 | (0.95; 1.07) | 1.07 | (0.92; 1.24) |
| **Residence** |  |  |  |  |
| Urban | 1.07 | (0.96; 1.20) | 1.06 | (0.79; 1.43) |
| Rural | 1.19 | (1.00; 1.43) | 1.19 | (1.00; 1.43) |
| Urban and rural mixed | 1.05 | (0.96; 1.20) | 1.13 | (1.00; 1.28) |
| **Publication language** |  |  |  |  |
| English | 1.04 | (0.99; 1.10) | 1.11 | (0.99; 1.24) |
| Chinese | 1.20 | (1.05; 1.36) | 1.21 | (0.91; 1.62) |

### Publication bias and quality assessment

Visual inspection of funnel plot and Egger’s regression test have been conducted to assess small-study effects or publication bias comparing famine births to pre- and post-famine births combined controls. Neither plot asymmetry (**Supplementary Figure 5**) nor significant Egger’s test was observed. The quality assessment covered three domains (sample, design, and analysis) with eight items for included studies (**Figure 5**). Each item can be scored as ‘good (2)’, ‘fair (1)’, and ‘poor (0)’, with a total score ranging 0-16. Quality of most studies was poor or moderate with a total score between 3-10 except four studies having a score of over 10 (Study # 3, 13, 14, 18).^27,35,50,58^ Most studies scored ‘good’ in outcome assessment but scored ‘fair’ or ‘poor’ in control selection and famine intensity assessment.

**Figure 5.**
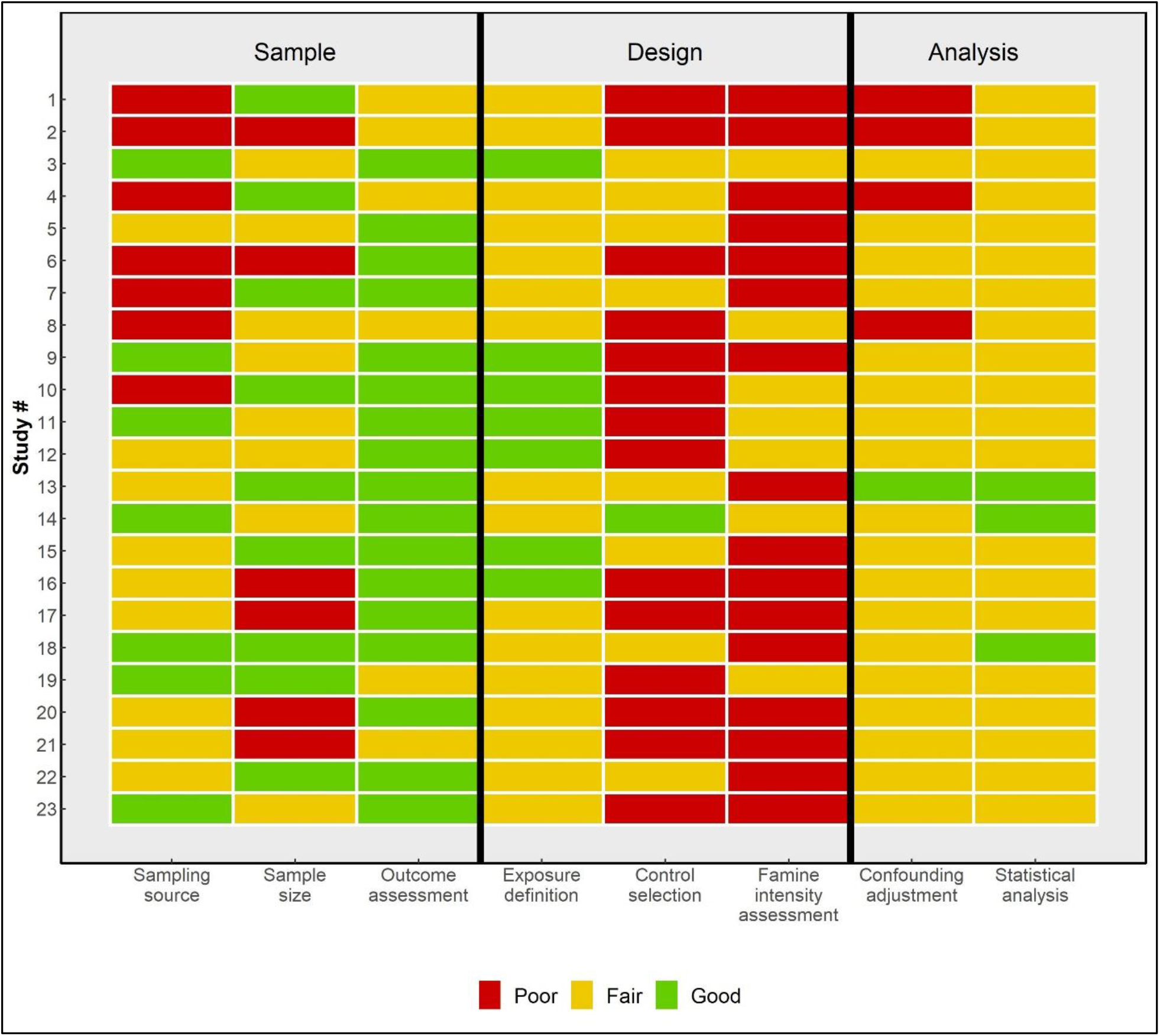
Quality assessment of included studies.

## DISCUSSION

In this study, we used meta-analysis as a comparative tool to examine how control selections can influence the results of Chinese famine studies on T2D and to identify study characteristics influencing study results. We found that the selection of control groups, including post-famine births, pre- and post-famine births combined, and pre-famine births, determines the direction of famine effects on T2D (negative, null, or protective). Existing studies predominantly used post-famine births as controls and are therefore subject to the bias caused by the age-difference between famine and post-famine births. This is because the risk of T2D increases with an increasing age among middle aged and older Chinese population. Studies with larger age differences between comparison groups had larger famine effects whichever control group is used. We also identified large variations and methodological problems in sampling method, famine intensity assessment, and confounding adjustment of included T2D studies. This adds difficulties to interpretation and comparison of results across studies.

While the number of original Chinese famine studies on T2D increased from eight^4^ to over 30 in the past five years, improvements in their quality are limited except for few studies (Study #13, 14, 18).^27,35,58^ Therefore, the key challenge is how methodological problems can be appropriately addressed to better quantify impacts of the famine in future studies. In the following paragraphs, we will, therefore, discuss main methodological problems identified through this systematic review and meta-analysis, and propose solutions to them. These problems include but not limit to: uncontrolled age difference, poorly assessed famine intensity, unsuitable sampling method, and inappropriate covariate adjustment.

First, the age effect on the development of T2D is strong, so it is important to address age differences between famine births and controls.^5,65^ Most recent studies continued to use post-famine births as controls and therefore mix age effects with famine effects. Their results have been widely interpreted as evidence that early-life famine exposure is a major risk factor for T2D and other adverse health conditions among populations in China and elsewhere.^3,14-17,19^ This bias cannot be fixed by adjusting for age in regression analysis because there is no overlap in the birth years of famine and post-famine births and T2D risk increases non-linearly with age.^5,23,66^ Using younger controls will always generate apparent ‘famine effects’ in older age groups (**Figure 3B and Supplementary Figure 2A**). Using pre- and post-famine births combined as controls, apparent famine effects will be substantially smaller (**Supplementary Figure 2B**). Using post-famine births as controls, famine effects can be even ‘protective’ (**Supplementary Figure 2C**). The larger age difference between comparison groups is, the larger effect estimate is likely to be (**Figure 4**). However, combining pre- and post-famine births will not always fix the problem because the age difference cannot be balanced out in some studies (**Figure 3A and 4**).^57,59,61^ This can lead to biased study results.

The famine affected all provinces of China, so it is difficult to find unexposed controls with a similar age to famine births.^67-69^ Fortunately, several analytical methods can be used to address this problem, including using ‘age-balanced’ controls, difference-in-difference (DID) models, and age-period-cohort (APC) approaches. Age-balanced controls can be created by combining pre- and post-famine births if the two groups had a similar magnitude of age difference compared to famine births and sample size of the two groups are comparable. DID models can also be used by including both time controls and place controls. In this method, famine births, pre-famine births, and post-famine births from areas of different famine intensity levels are compared. In regression models, an interaction term of time of birth and place of birth will be added to show when and where the famine effect is most pronounced.^70^ APC approaches can partition variation in T2D into age effects, period effects, and cohort effects, recognizing identification issues. There are successful examples of how these methods have been applied to Chinese famine studies.^34,35,71,72^ One shared characteristic of these improved analytical methods is that they all included pre-famine, famine, and post-famine births in areas with different levels of famine intensity.

Many Chinese famine studies also reported increased risk of T2D and other adverse health outcomes among pre-famine births comparing to post-famine births. It should be noted that age differences between these two groups were at least six years and can even be over ten years in some studies.^31,53,58,59^ Although pre-famine births have experienced famine in early childhood, no increased risk of T2D has been observed among pre-famine births in studies of other famines, including the Dutch and Ukraine famines.^73-75^ In the Chinese famine, the crude death increased from 10 to 25 per thousand (2.5-fold increase); in the Ukraine famine, the crude death rate increased from 8 to around 120 per thousand each year (15-fold increase).^69,76^ Furthermore, our exploration of CHARLS data suggested no increased risk of T2D over time among pre-famine births after taking age effect into consideration.^77^ Therefore, we expect no increased odds of T2D among pre-famine births caused by the Chinese famine.

Second, most of the included T2D studies did not carefully examine the intensity of the Chinese famine. This can lead to potential misclassifications of famine births and controls. Because individual levels of energy or food intake at the time of the famine are not available in Chinese famine studies, ecological data of the famine together with individuals’ birth information have been used to define famine exposure and assess exposure level.^4,78^ In Dutch and Ukraine famine studies, researchers established famine period and intensity using historical and demographic documents, and then compared ecological levels of exposure with individual birth information.^1,73,75^ However, most of the Chinese famine studies on health have not attempted to define famine exposure beyond the year and/or month of birth.^4,5^ In addition, inconsistencies in timing used to define the famine births and other groups were observed (**Supplementary Figure 1**). Given the nature of Chinese famine and lack of reliable administrative documents, it may never be possible to have as much relevant information as that in the Dutch famine studies.

Some studies did examine famine intensity, but methods used to assess intensity were problematic.^30,35,42,50,52-54^ Following a study by Luo et al. in 2006,^79^ Li et al. used mortality data to estimate famine intensity.^50^ An 50% increase in mortality rate comparing famine to pre-famine years was used as the cutoff point to classify famine intensity as ‘more severe’ vs. ‘less severe’ for each province.^50^ This method was also followed by later T2D studies^35,52,53^ and studies on many other health outcomes as well.^80-83^ It is unclear why this cutoff point was used, as it can lead to significant misclassifications of exposure levels. For example, the provinces of Jilin (56.4% increase in mortality), Guangdong (57.8% increase), and Anhui (474.9% increase) were all grouped together as ‘famine severe’ areas despite a large variation (i.e. nearly ten-fold) in increased mortality.^50,79^ This may bias effect estimates towards the null for famine births from extreme areas and away from the null for births from moderate areas.^23,66,84^ To better estimate a possible dose-response in famine effects, famine intensity should therefore be classified into more levels than two. Three or four levels will be more informative. Other studies^42,54^ have used grain productions to estimate famine severity, citing an economic study by Lin and Yang^85^ or using local grain data.^86^ Methods based on grain production can be problematic because the grain production alone in China was not the major cause of the famine.^67-69,85,87-99^ To assess famine intensity, documents and studies from additional disciplines should be examined, including history, demography, and economics. For example, our recent studies provided alternative methods to assess famine intensity at different regional levels.^77,100,101^ There is a need therefore to develop a robust famine intensity measurement to facilitate the identification of potential dose-response effects and the comparison of results across studies.

Third, most of the included T2D studies were secondary analyses of existing cross-sectional surveys or cohorts, none of which except for one^86^ was specifically designed to examine the impact of famine exposure on health outcomes (**Supplementary Table 2**). These cross-sectional surveys or cohorts may not be suitable to identify famine effects because their sampling methods were often problematic and key information for high-quality famine studies can be missing. For example, the convenience sampling used in multiple studies led to challenges in interpretation and limited generalizability of their findings because it is difficult to relate the study population to a well-defined population with and without famine exposure (**Supplementary Table 2**).^25,39,41,42,49,51,52^ When the sampling source is unclear, the study can be hardly salvaged even with appropriate control selection and robust famine intensity assessment.

Another problematic sampling issue is that some studies did not recruit pre-famine births at all, making it impossible to address the problem of age-difference.^39,54,56,66^ This is also true for the cohort specifically designed to examine famine effects.^54^ Besides, the population size of individuals born in famine years was usually much smaller than that of individuals born before and after the famine in most regions,^67,69^ which may compromise study power. Some key information was not necessarily collected in most studies, including place of birth and residence, familial socioeconomic status (SES) at the time of famine, and T2D biomarkers. Therefore, it is important to design studies that avoid these inherent problems of some existing surveys or cohorts. Pilot studies with oversampling of famine births are needed to identify areas or regions suitable for future famine studies.

Forth, most T2D studies adjusted for different sets of covariates. Some covariate adjustments, however, may not be appropriate. For example, it may be problematic to adjust for body size in estimating the association of famine exposure and T2D,^102^ because it remains unclear if body size is an effect modifier or a mediator of this relationship.^27,78,102-104^ If body size modifies the relationship between famine exposure and T2D, the analysis should be stratified by body size;^27,104^ if body size is a mediator of the relationship, there is no need to adjust for it.^78,103^ One way to address this question is to collect data for both T2D and body size over time and to examine how their interrelation may change over time and how changes in each are related to the other. Several studies adjusted for both body mass index (BMI) and waist circumference,^25,27,53,56^ which may lead to the problem of collinearity. Some studies, but not all, adjusted for rural vs. urban residence at the time of the study in view of a big rural-urban difference of T2D in China.^31,57^ It is also important to include the residence at birth in the analysis, because there was a substantial difference in famine severity in rural vs. urban areas during the famine. A potentially important factor that is often ignored is the familial SES at the time of famine, as this may both influence famine exposure^105,106^ and T2D.^107^ Familial SES at the time of famine has been shown to be important for health outcomes in Dutch famine studies.^70^

Some small studies adjusted for many covariates, forcing multivariate regressions on many empty cells. For example, Li et al. adjusted for six covariates with 17 cases of T2D among famine births.^50^ In several other studies, adjusted associations between famine exposure and T2D are very different from crude associations.^27,50,56,103^ For example, in one study, the crude OR is 1.25 (95% CI: 0.74-2.11) but an adjusted OR is 5.71 (95% CI: 1.53-21.2).^56^ Such differences in crude and adjusted estimates have seldom been explored but could lead to further insights.^103^ In the Dutch famine and Ukraine famine studies, adjusted associations agreed well with crude associations except for those adjusted for BMI.^73,78^ Confounding is a major challenge for causal inference in life-course epidemiologic studies.^108^ DAGs will be useful for future studies to examine the rationale for covariate adjustment.^109,110^

Methodological problems discussed above can also be observed in most Chinese famine studies on other health outcomes because similar data source and analytical methods are used. All too often, adverse health outcomes are linked to the early-life exposure of Chinese famine without proper consideration of these methodological problems.^4,5,66,100,101,103^ The universal use of post-famine births as controls in Chinese famine studies is the reason why most studies have reported negative famine effects on different health outcomes.

There are multiple studies of systematic review and/or meta-analysis examining the relationship between early-life famine exposure and health outcomes in adulthood.^4,6-12,111^ Most of these meta-analyses relied heavily on Chinese famine studies and aimed to ‘clarify’ the relationship between famine exposure and health outcomes, including T2D.^6,9,10^ These meta-analyses all reported a 1.4-fold increase in the risk for T2D after prenatal famine exposure,^6,9,10^ and we observed a similar summary estimate comparing famine births to post-famine births (**Supplementary Figure 2A**). We found that these meta-analyses directly pooled maximally adjusted effect estimates of prenatal famine exposure without assessing control selections in individual studies and without extracting tabular information on number of cases and population at risk. Therefore, they failed to recognize methodological problems discussed above, especially uncontrolled age differences between famine births and post-famine births in Chinese famine studies.^6-12^ This shows how meta-analysis of observational studies would generate misleading results unless careful examination of study methods is carried out.^21,22^ It is important to keep in mind using meta-analysis as an exploratory and comparative tool rather than a tool of producing ‘precise’ results especially for observational studies.

In contrast, our systematic review and meta-analysis of Chinese famines studies on T2D performed a careful examination of how control selections will influence study results. It demonstrated the relationship between age differences and effect estimates. It identified main methodological problems of uncontrolled age difference, poorly assessed famine intensity, unsuitable sampling method, and inappropriate covariate adjustment, and provided recommendations for future studies. Some limitations of this systematic review and meta-analysis need also to be acknowledged. Because of differences in the design and methods of included T2D studies, it may not be appropriate to use meta-analysis to calculate summary estimates. However, we used meta-analysis as a tool to examine systematic differences in controls used across studies as an important study characteristic influencing results.^23,47^ We also showed that effect estimates comparing famine births to pre- and post-famine births combined were stable after stratification by selected characteristics. As our study does not have access to the original data of most included studies, we are not able to examine some important questions. For example, it is unclear how adjusted associations could be so different from crude associations in some studies.^27,50,56,103^ Another question is that Dutch famine studies often showed increases in both glucose levels and body sizes among exposed individuals,^75,78^ but such patterns were not consistently observed in most Chinese famine studies.

We established that widely reported famine effects on T2D in China is driven by the selection of controls. The larger the age difference is, the larger the observed effect will be. We expect that the famine could have had an important impact on the risk and development of T2D among the Chinese population with prenatal famine exposure.^5,84^ Considering the current heavy burden of T2D in China, it is necessary to examine early-life environmental factors that may have contributed to this epidemic. Most of the current Chinese famine studies, however, have serious methodological shortcomings, including uncontrolled age difference, poorly assessed famine intensity, unsuitable sampling method, and inappropriate covariate adjustment. Better estimates of famine effects on T2D and other health conditions are needed by addressing these shortcomings. These efforts to improve the quality of Chinese famine studies will provide important evidence and recommendations for public health policy.

## Data Availability

All data produced in the present work are contained in the manuscript.

## Data sharing

The data used in this study are all presented in main tables or supplementary tables.

## Acknowledgements

We thank Drs. Steven Stellman, Henry Greenberg, Guohua Li, Hongwei Xu, and Silvia Martins for their comments and suggestions.

