## Supplementary materials for "Early-life exposure to the Chinese famine of 1959-61 and type 2 diabetes in adulthood: a systematic review and meta-analysis"

### Supplementary Text 1. Study protocol and search results

#### Section 1. Protocol

#### Objective

To conduct a systematic review and meta-analysis of the relationship between prenatal exposure to the Chinese famine of 1959-61 (Chinese famine) and adult type 2 diabetes (T2D)

#### Methods

The Preferred Reporting Items for Systematic Reviews and Meta-Analyses (PRISMA) and Meta-analysis of Observational Studies in Epidemiology (MOOSE) guidelines will be followed during all stages of the design, implementation, and reporting of this meta-analysis.

Definition of Exposure and Outcomes

1. Exposure: Prenatal exposure to the Chinese famine
2. Outcome: T2D, hyperglycemia, high blood sugar, and impaired fasting glucose in adulthood

#### Inclusion Criteria

1. Design and setting: Observational studies assessed the relationship between prenatal exposure to the Chinese famine and diabetes or blood glucose dysregulation. Commentaries, protocols, or review articles will be included as a source of references.
2. Study population: Main study population included adult populations in China who were born around the 1950-60s (before, during, and after the Chinese famine).
3. Exposure: The Chinese famine of 1959-61 was defined as a prenatal exposure or risk factor.
4. Outcome: Health outcomes had been assessed with comparable methods for different birth groups with and without prenatal exposure to the Chinese famine.
    a. T2D measured by different assessment criteria
    b. Blood glucose level, hyperglycemia, impaired fasting glucose
5. Effect measure: Studies provided an estimate of the difference in the outcome specified and a measure of uncertainty for the reported difference, or report the relative risk estimates (or odds ratio) for other outcomes with standard error (or information to compute them; or such data can be obtained from authors).
6. Language: English and Chinese

**Exclusion Criteria**

1. Studies did not provide clear descriptions of the study population, study design, analytical method, and effect estimate.
2. When several studies were based on the same or overlapping cohorts reporting related results, the study that had the most comprehensive information on exposure definition, outcome, and effect measure will be used as a representative study, and the others will be excluded.

**Databases**

1. First broad search:

English databases

   a. PubMed/MEDLINE
   b. Embase (Biomedical database from Elsevier)
   c. Web of Science

Chinese databases

   a. Chinese Wanfang Data
   b. Chinese National Knowledge Infrastructure (CNKI)

2. Second search:

   a. Additional online searches through Google Scholar and ResearchGate
   b. Hand searches of references of related studies

**Section 2 Search Terms and Results**

<u>English databases</u>

PubMed/MEDLINE: searched conducted on Feb 8$^{th}$, 2022 and 9,382 items identified
(China OR Chinese) AND (famine OR undernutrition OR starvation OR malnutrition) OR great leap forward OR great famine

Embase: searched conducted on Feb 8$^{th}$, 2022 and 12,612 items identified
(china OR chinese) AND (famine OR undernutrition OR starvation OR malnutrition) OR (great AND leap AND forward) OR (great AND famine)

Web of Science: searched conducted on Feb 8$^{th}$, 2022 and 10,844 items identified

(China OR Chinese) AND (famine OR undernutrition OR starvation OR malnutrition) OR great leap forward OR great famine

<u>Chinese databases</u>

Chinese Wanfang Data: searched conducted on Feb 8th, 2022 and 11,768 items identified

中国饥荒 OR 大跃进 OR 三年自然灾害

Chinese National Knowledge Infrastructure: searched conducted on Feb 8th, 2022 and 3,093 items identified

中国饥荒 OR 大跃进 OR 三年自然灾害

<u>Other sources</u>

Commentaries, protocols, review articles, cited references, and Google Scholar and ResearchGate: 10 identified

Total from preliminary search: 47,709

**Information Extracted from Eligible Studies**

a. Author and publication information: Authors, year of publication, publication language, journal, study title
b. Study characteristics: Data source, study design, sampling method, sample size, exposure definition, control selection, outcome measurement, famine intensity assessment, analytical method, covariate adjustment, reported result
c. Time windows used to define different comparison groups and their mean ages
d. Tabular information: Numbers of T2D cases and populations at risk for different comparison groups (fetal exposed group and control group) with and without stratification by selected characteristics, including sex, mean age at the survey, outcome measurement, reported famine intensity, residence, publication language

### Supplementary Text 2. Quality Assessment Coding Criteria

For each main study characteristic, we assigned a score of ‘good (2)’, ‘fair (1)’, and ‘poor (0)’. The modified criteria are described as bellow.

**Sampling source**

Good: the study population can be related to a well-defined population with and without famine exposure and different levels of famine exposure, e.g., national cohort studies based on systematic sampling.

Fair: the study population can be related to a well-defined population with and without famine exposure but not necessarily different levels of famine exposure, e.g., prefecture-level cohort studies based on systematic sampling.

Poor: the study population cannot reflect the disease pattern among individuals with and without famine exposure, e.g., hospital-based studies based on convenience sampling.

**Sample size**

Most Chinese famine studies has a prevalence of T2D between 5% to 15%.

Good: the monthly number of exposed subjects is over 40, which in turn will have at least 72 T2D cases among subjects born in 1959-61.

Fair: the monthly number of exposed subjects is between 10 and 40, which in turn will have at least 18 T2D cases among subjects born in 1959-61.

Poor: the monthly number of exposed subjects is below 10 or not reported or cannot be calculated.

**Outcome assessment**

Good: the outcome of T2D or glucose dysregulation is assessed clinically or with similar standards.

Fair: the outcome of T2D or glucose dysregulation is assessed but not clinically.

Poor: the outcome assessment method is not clearly described.

**Exposure definition**

Good: the famine exposure is defined by date of year for the famine period with adequate explanation or discussion.
Fair: the exposure is defined by date of year for the famine period with some explanation or discussion.
Poor: the exposure definition is not clearly described or justified.

**Control selection**
Good: both appropriate time controls and place controls were recruited.
Fair: either appropriate time controls or place controls were recruited.
Poor: no appropriate controls were recruited.

**Famine intensity assessment**
Good: appropriate and different data sources are used to evaluate famine intensity, e.g., census data and mortality data to quantify famine intensity.
Fair: single data source is used to evaluate famine intensity, e.g., morality data with a 50% excess death rate.
Poor: no data is used or reported to evaluate famine intensity.

**Confounding adjustment**
Good: the confounding adjustment is conducted with adequate explanation or discussion.
Fair: the confounding adjustment is conducted with some explanation or discussion.
Poor: the confounding adjustment is not conducted or reported.

**Statistical analysis**
Good: appropriate statistical analysis is conducted and supplemented by sensitivity analysis.
Fair: appropriate statistical analysis is conducted.
Poor: the statistical analysis is not properly conducted or clearly reported.

**Supplementary Table 1. PRISMA checklist of items to include when reporting a systematic review and meta-analysis**

| Section and Topic | Item # | Checklist item | Location where item is reported |
|---|---|---|---|
| **TITLE** | | | |
| Title | 1 | Identify the report as a systematic review. | 1 |
| **ABSTRACT** | | | |
| Abstract | 2 | See the PRISMA 2020 for Abstracts checklist. | 1 |
| **INTRODUCTION** | | | |
| Rationale | 3 | Describe the rationale for the review in the context of existing knowledge. | 2-3 |
| Objectives | 4 | Provide an explicit statement of the objective(s) or question(s) the review addresses. | 2-3 |
| **METHODS** | | | |
| Eligibility criteria | 5 | Specify the inclusion and exclusion criteria for the review and how studies were grouped for the syntheses. | 3 |
| Information sources | 6 | Specify all databases, registers, websites, organisations, reference lists and other sources searched or consulted to identify studies. Specify the date when each source was last searched or consulted. | 3 |
| Search strategy | 7 | Present the full search strategies for all databases, registers and websites, including any filters and limits used. | 3 and Supplementary Text 1 |
| Selection process | 8 | Specify the methods used to decide whether a study met the inclusion criteria of the review, including how many reviewers screened each record and each report retrieved, whether they worked independently, and if applicable, details of automation tools used in the process. | 4 |

| Section and Topic | Item # | Checklist item | Location where item is reported |
|---|---|---|---|
| Data collection process | 9 | Specify the methods used to collect data from reports, including how many reviewers collected data from each report, whether they worked independently, any processes for obtaining or confirming data from study investigators, and if applicable, details of automation tools used in the process. | 5 |
| Data items | 10a | List and define all outcomes for which data were sought. Specify whether all results that were compatible with each outcome domain in each study were sought (e.g. for all measures, time points, analyses), and if not, the methods used to decide which results to collect. | 5 |
| | 10b | List and define all other variables for which data were sought (e.g. participant and intervention characteristics, funding sources). Describe any assumptions made about any missing or unclear information. | 5 |
| Study risk of bias assessment | 11 | Specify the methods used to assess risk of bias in the included studies, including details of the tool(s) used, how many reviewers assessed each study and whether they worked independently, and if applicable, details of automation tools used in the process. | 4 |
| Effect measures | 12 | Specify for each outcome the effect measure(s) (e.g. risk ratio, mean difference) used in the synthesis or presentation of results. | 4 |
| Synthesis methods | 13a | Describe the processes used to decide which studies were eligible for each synthesis (e.g. tabulating the study intervention characteristics and comparing against the planned groups for each synthesis (item #5)). | 4-5 |
| | 13b | Describe any methods required to prepare the data for presentation or synthesis, such as handling of missing summary statistics, or data conversions. | 4-5 |
| | 13c | Describe any methods used to tabulate or visually display results of individual studies and syntheses. | 4-5 |

| Section and Topic | Item # | Checklist item | Location where item is reported |
|---|---|---|---|
| | 13d | Describe any methods used to synthesize results and provide a rationale for the choice(s). If meta-analysis was performed, describe the model(s), method(s) to identify the presence and extent of statistical heterogeneity, and software package(s) used. | 4-5 |
| | 13e | Describe any methods used to explore possible causes of heterogeneity among study results (e.g. subgroup analysis, meta-regression). | 4-5 |
| | 13f | Describe any sensitivity analyses conducted to assess robustness of the synthesized results. | 4-5 |
| Reporting bias assessment | 14 | Describe any methods used to assess risk of bias due to missing results in a synthesis (arising from reporting biases). | 5 |
| Certainty assessment | 15 | Describe any methods used to assess certainty (or confidence) in the body of evidence for an outcome. | 4 |
| **RESULTS** | | | |
| Study selection | 16a | Describe the results of the search and selection process, from the number of records identified in the search to the number of studies included in the review, ideally using a flow diagram. | 6 |
| | 16b | Cite studies that might appear to meet the inclusion criteria, but which were excluded, and explain why they were excluded. | 3 and 6 |
| Study characteristics | 17 | Cite each included study and present its characteristics. | 5 |
| Risk of bias in studies | 18 | Present assessments of risk of bias for each included study. | 7 |

| Section and Topic | Item # | Checklist item | Location where item is reported |
|---|---|---|---|
| Results of individual studies | 19 | For all outcomes, present, for each study: (a) summary statistics for each group (where appropriate) and (b) an effect estimate and its precision (e.g. confidence/credible interval), ideally using structured tables or plots. | 6-7 |
| Results of syntheses | 20a | For each synthesis, briefly summarise the characteristics and risk of bias among contributing studies. | 7 |
| | 20b | Present results of all statistical syntheses conducted. If meta-analysis was done, present for each the summary estimate and its precision (e.g. confidence/credible interval) and measures of statistical heterogeneity. If comparing groups, describe the direction of the effect. | 6-7 |
| | 20c | Present results of all investigations of possible causes of heterogeneity among study results. | 6-7 |
| | 20d | Present results of all sensitivity analyses conducted to assess the robustness of the synthesized results. | 6-7 |
| Reporting biases | 21 | Present assessments of risk of bias due to missing results (arising from reporting biases) for each synthesis assessed. | 7 |
| Certainty of evidence | 22 | Present assessments of certainty (or confidence) in the body of evidence for each outcome assessed. | 6-7 |
| **DISCUSSION** | | | |
| Discussion | 23a | Provide a general interpretation of the results in the context of other evidence. | 7-8 |
| | 23b | Discuss any limitations of the evidence included in the review. | 7-11 |
| | 23c | Discuss any limitations of the review processes used. | 12 |
| | 23d | Discuss implications of the results for practice, policy, and future research. | 12-13 |
| **OTHER INFORMATION** | | | |

| Section and Topic | Item # | Checklist item | Location where item is reported |
|---|---|---|---|
| Registration and protocol | 24a | Provide registration information for the review, including register name and registration number, or state that the review was not registered. | 5 |
| | 24b | Indicate where the review protocol can be accessed, or state that a protocol was not prepared. | 5 |
| | 24c | Describe and explain any amendments to information provided at registration or in the protocol. | 5 |
| Support | 25 | Describe sources of financial or non-financial support for the review, and the role of the funders or sponsors in the review. | NA |
| Competing interests | 26 | Declare any competing interests of review authors. | NA |
| Availability of data, code and other materials | 27 | Report which of the following are publicly available and where they can be found: template data collection forms; data extracted from included studies; data used for all analyses; analytic code; any other materials used in the review. | NA |

**Supplementary Table 2. Additional characteristics of included Chinese famine studies on T2D**

| Study # | Authors | Study design | Sampling method | Sample size* | Fetal exposed | Controls | Famine intensity measurement | Main statistical method | Covariate adjustment |
|---|---|---|---|---|---|---|---|---|---|
| 1 | Liu et al. 2009 | Cross-sectional | Hospital based convenience sampling | 4,640 | 1,468 | 3,172 | None | Chi-square test | None |
| 2 | Guan et al. 2009 | Cross-sectional | Hospital based random sampling of Chongqing Gangtie Group | 293 | 74 | 84 | None | Logistic regression | Sex, occupation, education, smoking, drinking, physical activity, family disease history, stress, birth weight, diet, and BMI |
| 3 | Li et al. 2010 | Cross-sectional | National systematic sampling | 7,874 | 1,005 | 1,954 | 50% excess death rate | Logistic regression | Sex, education, smoking, drinking, physical activity, family history of diabetes |
| 4 | Li et al. 2010 | Cross-sectional | Hospital based convenience sampling | 10,426 | 2,425 | 3,261 | None | T-test and Chi-square test | None |
| 5 | Zhang et al. 2010 | Cross-sectional | Prefecture systematic sampling | 949 | 461 | 488 | None | Logistic regression | Sex, age, smoking, drinking, and BMI |
| 6 | Zhao et al. 2013 | Cross-sectional | Hospital based convenience sampling | 847 | 91 | 399 | None | Logistic regression | Sex, education, smoking, physical activity, family history of diabetes, region, and diet |
| 7 | Li et al. 2014 | Cross-sectional | Corporation based convenience sampling | 19,347 | 3,314 | 12,043 | None | Logistic regression | Sex, age, education, income, smoking, drinking, physical activity, BMI, WHR, SBP DBP, TC, TG, LDL-C, and HDL-C |
| 8 | Zhang et al. 2014 | Cross-sectional | Hospital based convenience sampling | 4,212 | 1,233 | 1,999 | Grain production | T-test | None |

| | | | | | | | | | |
|---|---|---|---|---|---|---|---|---|---|
| | | | | | | | between 1953 and 1962 | | |
| 9 | Wang et al. 2015 | Cross-sectional | Multi-province systematic sampling | 6,897 | 745 | 1,808 | None | Logistic regression | Sex, age, waist circumference, height, rural/urban residence, and economic development |
| 10 | Wang et al. 2016 | Cross-sectional | Corporation based convenience sampling | 7,801 | 1,266 | 938 | 50% excess death rate | Logistic regression | Sex, famine severity, smoking, drinking, metabolic equivalent, family history of diabetes, and BMI |
| 11 | Wang et al. 2017 | Cross-sectional | Multi-province systematic sampling | 3,973 | 489 | 1,632 | 50% excess death rate | Logistic regression | Sex, age, education, smoking, and waist circumference |
| 12 | Li et al. 2017 | Cross-sectional | County systematic sampling | 2,068 | 983 | 1,085 | Grain production between 1953 and 1962 | Logistic regression | Sex, age, smoking, physical activity, diet, and BMI |
| 13 | Meng et al. 2018 | Longitudinal | National volunteer-based sampling | 88,830 | 18,879 | 69,951 | None | Cox-proportional hazards regression | Sex, age, education, marital status, smoking, drinking, physical activity, family history of diabetes, menopausal status, diet, WHR and BMI |
| 14 | Wang et al. 2018 | Cross-sectional | National systematic sampling | 4,138 | 832 | 3,306 | 50% excess death rate | Logistic regression | Sex, education, smoking, drinking, physical activity, and BMI |
| 15 | Zhang et al. 2018 | Cross-sectional | Province based systematic sampling | 5,960 | 1,442 | 1,986 | None | Logistic regression | Education, region, smoking, drinking, physical activity, fruit intake, and BMI |

| | | | | | | | | | |
|---|---|---|---|---|---|---|---|---|---|
| 16 | Zhou et al. 2018 | Cross-sectional | Prefecture systematic sampling | 939 | 84 | 381 | None | Quantile regression | Sex, education, income, rural/urban residence, smoking, drinking, physical activity, and family disease history |
| 17 | Liu et al. 2019 | Cross-sectional | County systematic sampling | 511 | 177 | 334 | None | Logistic regression | Age, education, residence, family history of diabetes, SBP, DBP obesity, and BMI |
| 18 | Lu et al. 2020 | Longitudinal | National multi-center sampling | 77,925 | 13,195 | 23,582 | None | Relative risk regression | Sex, age, family history of diabetes, drinking, education, and ideal cardiovascular health metrics |
| 19 | Zhang et al. 2020 | Cross-sectional | National systematic sampling | 7830 | 4081 | 3749 | 50% excess death rate | Logistic regression | Sex, economic status, education level, physical exercise, sedentary time, smoking, drinking, dietary factors, and BMI |
| 20 | Qi et al. 2020 | Cross-sectional | County cluster sampling | 8868 | NR | NR | None | Logistic regression | Age, smoking, drinking, physical exercise, family history of diabetes, education, marital status, and BMI |
| 21 | Ning et al. 2021 | Cross-sectional | County cluster sampling | 3418 | 213 | 1407 | None | Logistic regression | Age, family history of diabetes, BMI, residential areas, education, income level, total cholesterol, occupation, physical activity, smoking and drinking |
| 22 | Zhang et al. 2022 | Cross-sectional | Multiple hospital-based medical records | 361,639 | NR | NR | None | Linear regression | Sex |

| | | | | | | | | | |
|---|---|---|---|---|---|---|---|---|---|
| 23 | Huo et al. 2022 | Cross-sectional | Province based systematic sampling | 11,640 | 1,153 | 3,162 | None | Logistic regression | Age, sex, education, marital status, average monthly income, smoking, drinking, physical activity, BMI, high-fat diet, fruit-vegetable intake, family history of diabetes, and air pollutants |

* The sample included pre-famine, famine, and post-famine births. Some studies did not include pre-famine births.

NR: not reported.

**Supplementary Table 3. Mean age at the survey for different comparison groups**

| Study number | Average age (in years) | | | |
|---|---|---|---|---|
| | Pre-famine births | Famine births | Post-famine births | Pre- and post-famine births |
| **1** | 43.5 | 40.5 | 37.5 | 40.5 |
| **2** | - | - | - | - |
| **3** | 46.5 | 41.5 | 38.5 | 41.3 |
| **4** | - | - | - | - |
| **5** | - | - | 48.3 | 48.3 |
| **6** | 55.0 | 52.0 | 49.0 | 52.8 |
| **7** | - | 46.4 | - | 46.3 |
| **8** | - | - | - | - |
| **9** | 60.5 | 53.5 | 45.5 | 53.2 |
| **10** | 56.1 | 53.1 | 50.0 | 54.1 |
| **11** | 61.4 | 53.5 | 42.8 | 54.5 |
| **12** | - | 52.0 | 49.0 | - |
| **13** | 48.5 | 45.6 | 42.5 | 45.2 |
| **14** | 54.0 | 51.8 | 48.8 | 52.2 |
| **15** | 54.5 | 51.6 | 47.7 | 51.0 |
| **16** | 54.6 | 51.6 | 48.6 | 51.5 |
| **17** | - | 55.8 | 52.1 | - |
| **18** | 57.3 | 50.6 | 44.8 | 52.7 |
| **19** | - | 50.9 | 48.4 | - |
| **20** | - | - | - | - |
| **21** | 54.1 | 47.9 | 41.2 | 46.6 |
| **22** | - | - | - | - |
| **23** | 61.4 | 56.2 | 53.1 | 58.9 |

**Supplementary Figure 1. Exposure definition timing in Chinese famine studies.**

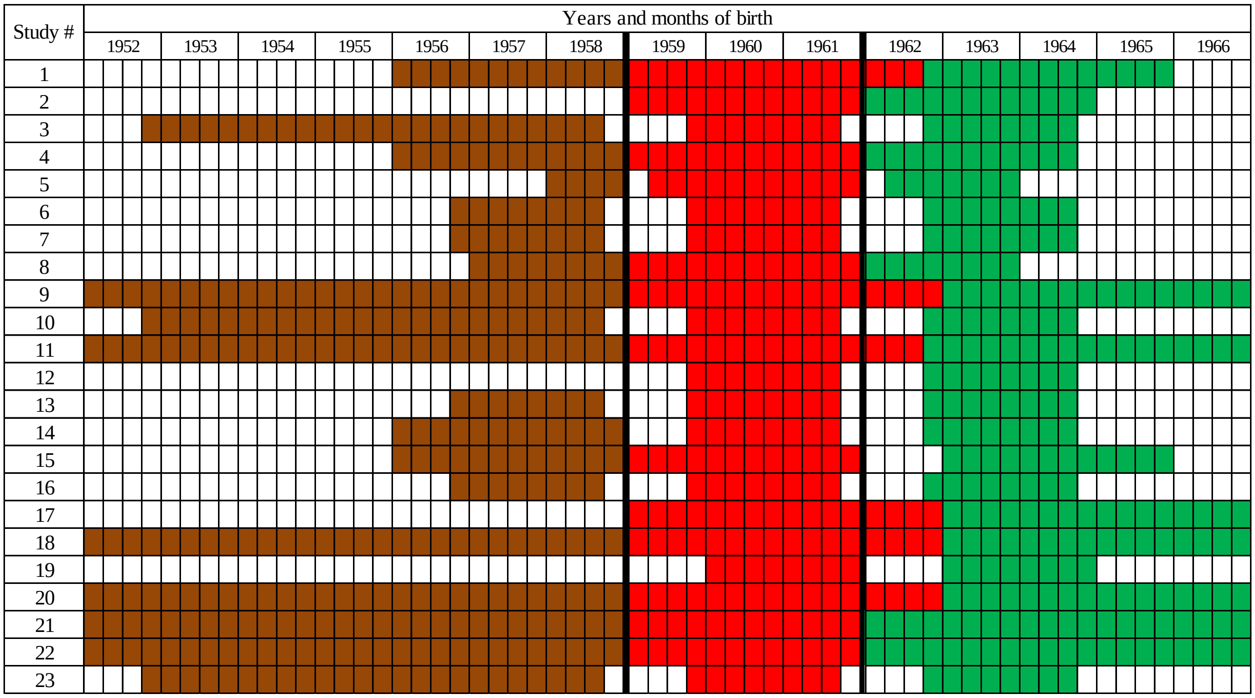


* Brown: pre-famine births; red: famine births; green: post-famine births.

**Supplementary Figure 2A. Effect estimates of famine exposure on T2D comparing famine births with post-famine births**

| Study # Authors | Famine T2D+ | Famine Total | Post-famine T2D+ | Post-famine Total | OR | 95% CI | Weight |
|---|---|---|---|---|---|---|---|
| 1. Liu et al. 2007 | 45 | 1468 | 74 | 3172 | 1.32 | [0.91; 1.93] | 5.3% |
| 2. Guan et al. 2009 | . | . | . | . | | | 0.0% |
| 3. Li et al. 2010 | 17 | 1005 | 31 | 1954 | 1.07 | [0.59; 1.94] | 2.8% |
| 4. Li et al. 2010 | . | . | . | . | | | 0.0% |
| 5. Zhang et al. 2010 | 51 | 461 | . | . | | | 0.0% |
| 6. Zhao et al. 2013 | 7 | 91 | 34 | 399 | 0.89 | [0.38; 2.09] | 1.5% |
| 7. Li et al. 2014 | 298 | 3314 | . | . | | | 0.0% |
| 8. Zhang et al. 2014 | . | . | . | . | | | 0.0% |
| 9. Wang et al. 2015 | 96 | 745 | 132 | 1808 | 1.88 | [1.42; 2.48] | 7.2% |
| 10. Wang et al. 2016 | 145 | 1266 | 111 | 938 | 0.96 | [0.74; 1.25] | 7.5% |
| 11. Wang et al. 2017 | 81 | 489 | 133 | 1632 | 2.24 | [1.66; 3.01] | 6.7% |
| 12. Li et al. 2017 | 110 | 983 | 61 | 1085 | 2.12 | [1.53; 2.93] | 6.2% |
| 13. Meng et al. 2018 | 318 | 138096 | 498 | 288237 | 1.33 | [1.16; 1.54] | 10.6% |
| 14. Wang et al. 2018 | 113 | 832 | 139 | 1536 | 1.58 | [1.21; 2.06] | 7.5% |
| 15. Zhang et al. 2018 | 122 | 1442 | 115 | 1986 | 1.50 | [1.15; 1.96] | 7.5% |
| 16. Zhou et al. 2018 | . | . | . | . | | | 0.0% |
| 17. Liu et al. 2019 | 27 | 177 | 42 | 334 | 1.25 | [0.74; 2.11] | 3.4% |
| 18. Lu et al. 2020 | 792 | 13195 | 978 | 23582 | 1.48 | [1.34; 1.62] | 11.7% |
| 19. Zhang et al. 2020 | 262 | 4081 | 193 | 3749 | 1.26 | [1.04; 1.53] | 9.3% |
| 20. Qi et al. 2020 | . | . | . | . | | | 0.0% |
| 21. Ning et al. 2021 | 27 | 213 | 76 | 1407 | 2.54 | [1.60; 4.05] | 4.0% |
| 22. Zhang et al. 2022 | . | . | . | . | | | 0.0% |
| 23. Huo et al. 2022 | 157 | 1153 | 285 | 3162 | 1.59 | [1.29; 1.96] | 8.9% |
| **Fixed effect model** | | **169011** | | **334981** | **1.46** | **[1.38; 1.55]** | **--** |
| **Random effects model** | | | | | **1.50** | **[1.34; 1.68]** | **100.0%** |

Heterogeneity: $I^2 = 64\%$, $\tau^2 = 0.0258$, $p < 0.01$

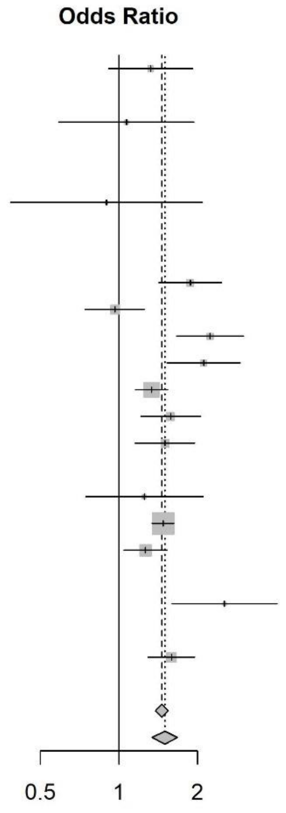


Boxes and horizontal lines represent odds ratios and 95% confidence intervals (CIs). The size of each box is proportional to the weight of the report for each outcome. Diamonds represent the 95% CI for pooled estimates of effect and are centered on the pooled odds ratio of a fixed-effect model or a random-effects model.

**Supplementary Figure 2B. Effect estimates of famine exposure on T2D comparing famine births with pre- and post-famine births combined**

| Study # | Famine | | Combined | | | | |
|---|---|---|---|---|---|---|---|
| Authors | T2DM+ | Total | T2DM+ | Total | OR | 95% CI | Weight |
| 1. Liu et al. 2007 | 45 | 1468 | . | . | | | 0.0% |
| 2. Guan et al. 2009 | . | . | . | . | | | 0.0% |
| 3. Li et al. 2010 | 17 | 1005 | 77 | 3608 | 0.79 | [0.46; 1.34] | 2.9% |
| 4. Li et al. 2010 | . | . | . | . | | | 0.0% |
| 5. Zhang et al. 2010 | 51 | 461 | 35 | 488 | 1.61 | [1.03; 2.53] | 3.7% |
| 6. Zhao et al. 2013 | 7 | 91 | 61 | 575 | 0.70 | [0.31; 1.59] | 1.4% |
| 7. Li et al. 2014 | 298 | 3314 | 928 | 12043 | 1.18 | [1.03; 1.36] | 11.5% |
| 8. Zhang et al. 2014 | . | . | . | . | | | 0.0% |
| 9. Wang et al. 2015 | 96 | 745 | 443 | 3719 | 1.09 | [0.86; 1.39] | 8.1% |
| 10. Wang et al. 2016 | 145 | 1266 | 387 | 2870 | 0.83 | [0.68; 1.02] | 9.1% |
| 11. Wang et al. 2017 | 81 | 489 | 540 | 3478 | 1.08 | [0.84; 1.39] | 7.5% |
| 12. Li et al. 2017 | 110 | 983 | . | . | | | 0.0% |
| 13. Meng et al. 2018 | 318 | 138096 | 1054 | 515731 | 1.13 | [0.99; 1.28] | 11.9% |
| 14. Wang et al. 2018 | 113 | 832 | 340 | 3306 | 1.37 | [1.09; 1.72] | 8.3% |
| 15. Zhang et al. 2018 | 122 | 1442 | 238 | 3568 | 1.29 | [1.03; 1.62] | 8.3% |
| 16. Zhou et al. 2018 | . | . | . | . | | | 0.0% |
| 17. Liu et al. 2019 | 27 | 177 | . | . | | | 0.0% |
| 18. Lu et al. 2020 | 792 | 13195 | 4050 | 64730 | 0.96 | [0.88; 1.03] | 13.4% |
| 19. Zhang et al. 2020 | 262 | 4081 | . | . | | | 0.0% |
| 20. Qi et al. 2020 | . | . | . | . | | | 0.0% |
| 21. Ning et al. 2021 | 27 | 213 | 190 | 2418 | 1.70 | [1.11; 2.62] | 4.0% |
| 22. Zhang et al. 2022 | . | . | . | . | | | 0.0% |
| 23. Huo et al. 2022 | 157 | 1153 | 1224 | 10487 | 1.19 | [1.00; 1.43] | 10.0% |
| **Fixed effect model** | | **169011** | | **627021** | **1.06** | **[1.01; 1.12]** | **--** |
| **Random effects model** | | | | | **1.12** | **[1.02; 1.24]** | **100.0%** |

Heterogeneity: $I^2 = 66\%$, $\tau^2 = 0.0179$, $p < 0.01$

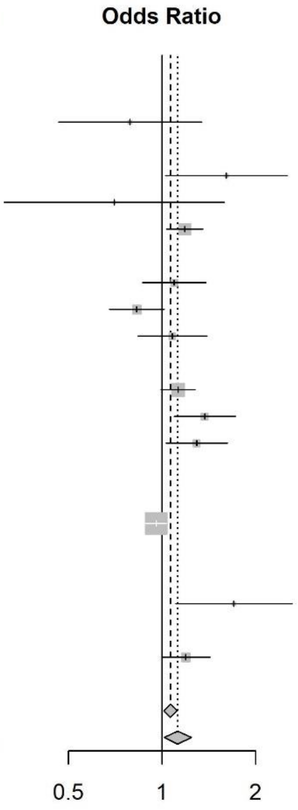


Boxes and horizontal lines represent odds ratios and 95% confidence intervals (CIs). The size of each box is proportional to the weight of the report for each outcome. Diamonds represent the 95% CI for pooled estimates of effect and are centered on the pooled odds ratio of a fixed-effect model or a random-effects model.

**Supplementary Figure 2C. Effect estimates of famine exposure on T2D comparing famine births with pre-famine births**

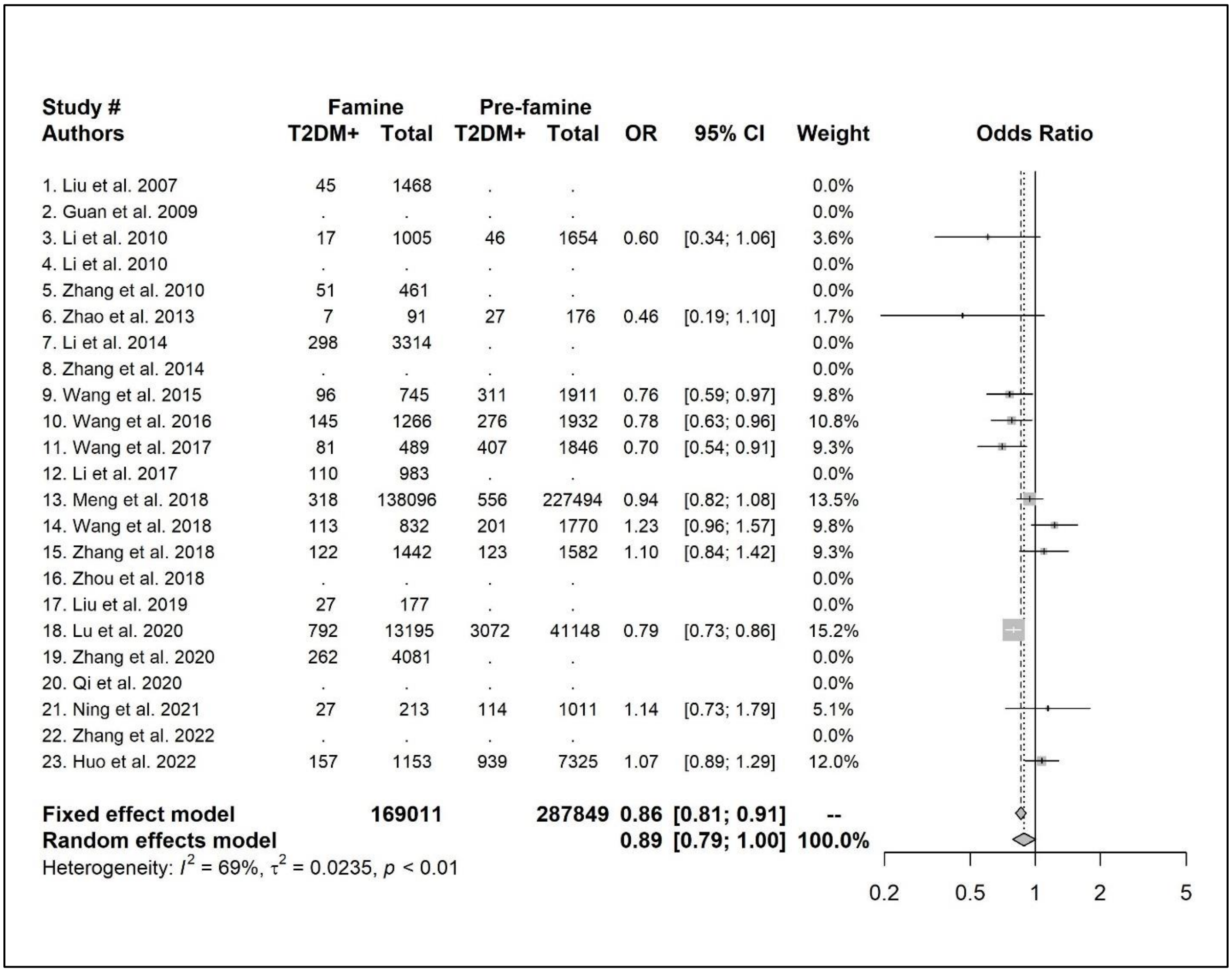


Boxes and horizontal lines represent odds ratios and 95% confidence intervals (CIs). The size of each box is proportional to the weight of the report for each outcome. Diamonds represent the 95% CI for pooled estimates of effect and are centered on the pooled odds ratio of a fixed-effect model or a random-effects model.

**Supplementary Figure 3A. Effect estimates of leave-one-out analysis comparing famine births with post-famine births**

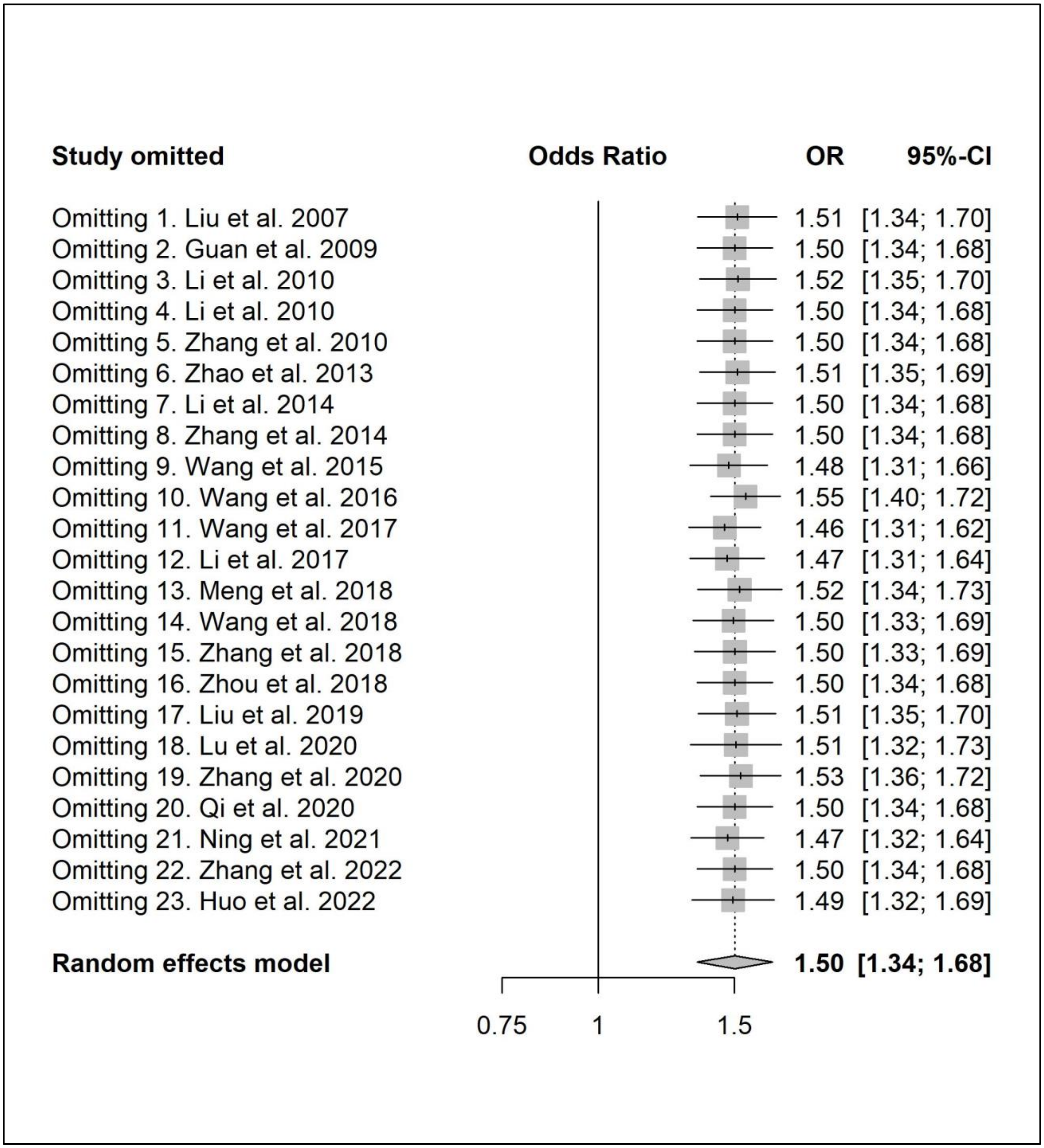

**Supplementary Figure 3B. Effect estimates of leave-one-out analysis comparing famine births with pre- and post-famine births combined**

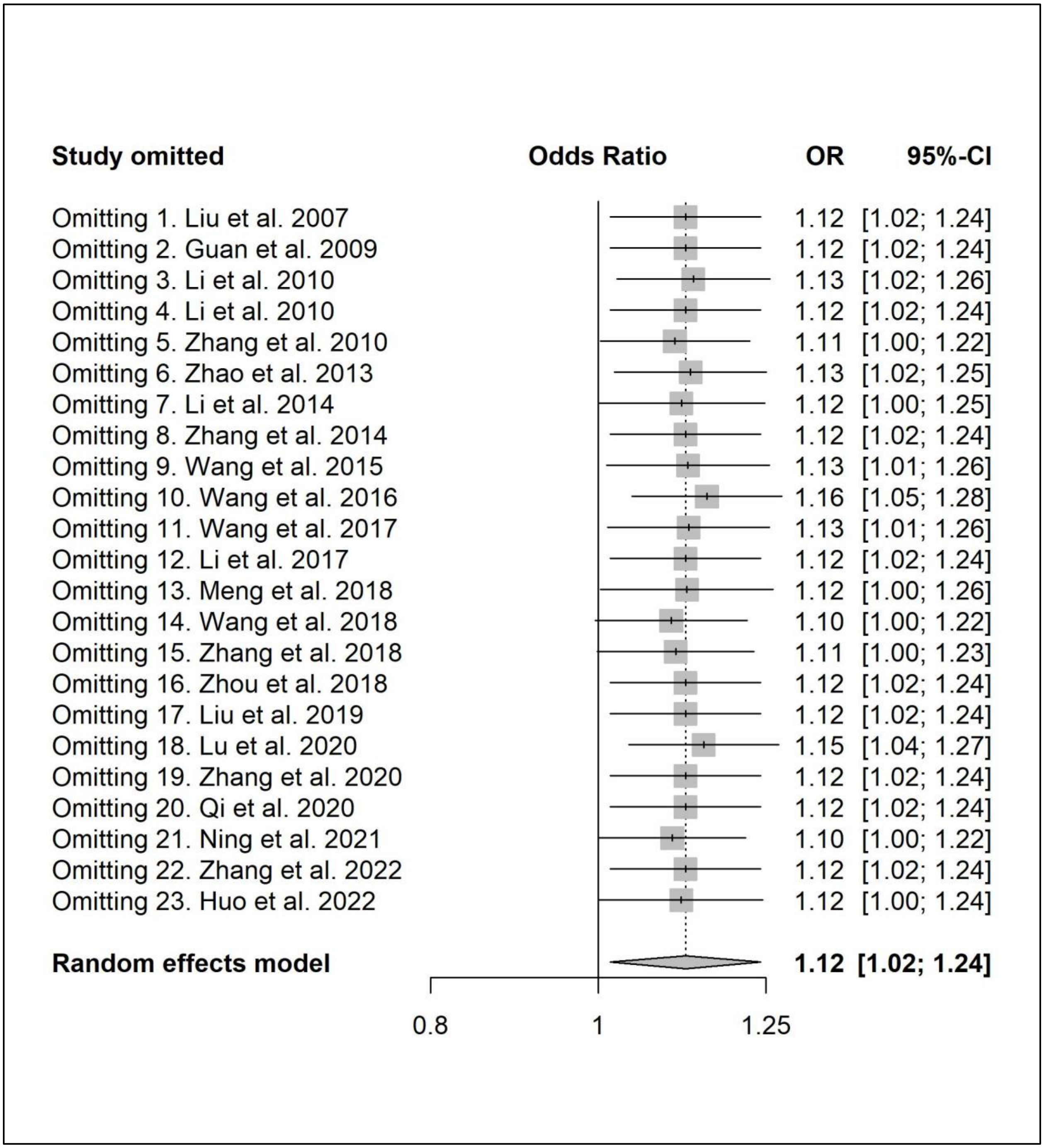

**Supplementary Figure 3C. Effect estimates of leave-one-out analysis comparing famine births with pre-famine births**

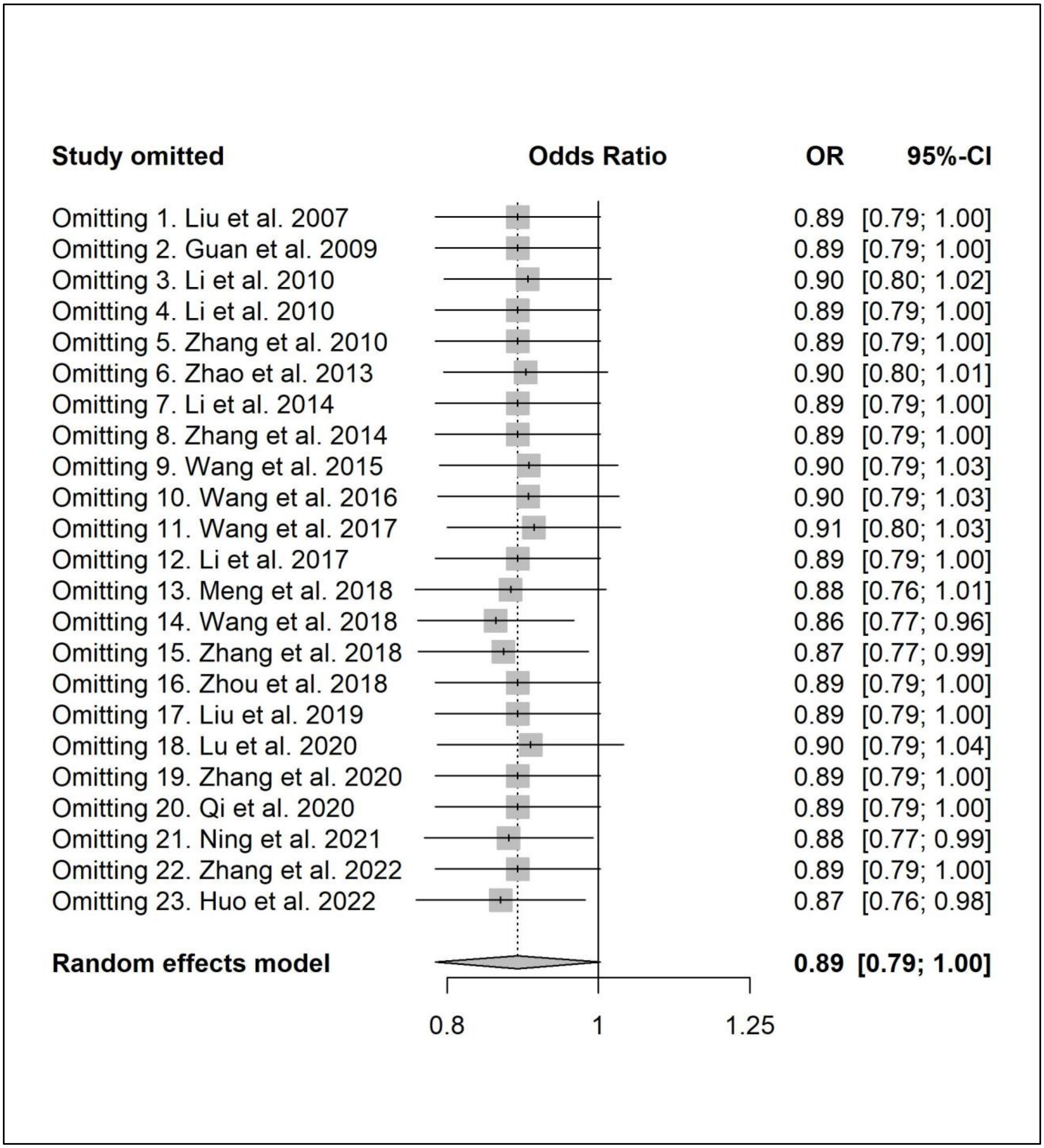

**Supplementary Figure 4A. Effect estimates of famine exposure on T2D comparing famine births with pre- and post-famine births combined after stratification by sex**

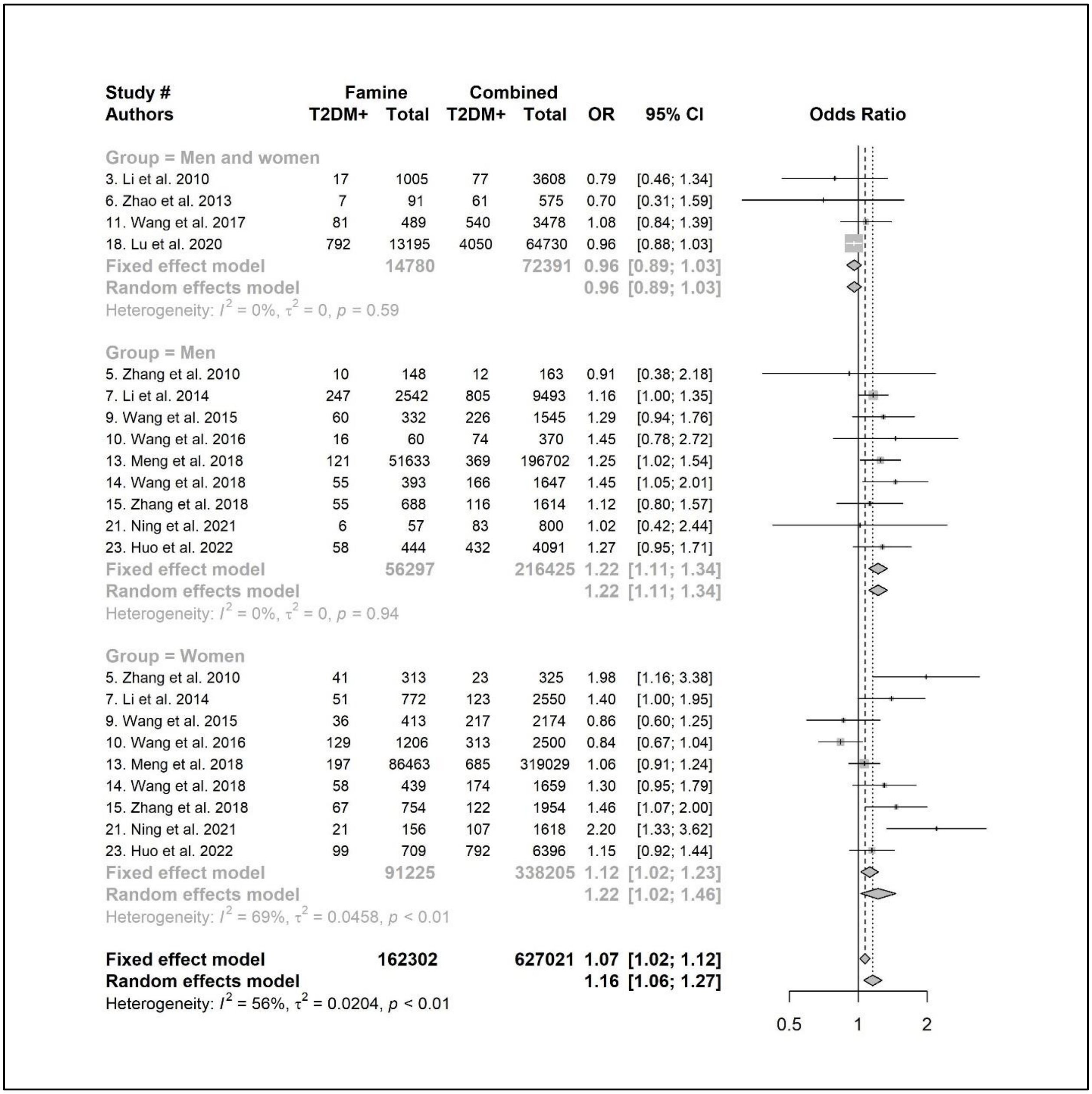


* The total number of famine births is 162,302 in the subgroup analysis of Supplementary Figure 4A-F, which is smaller than the total number of famine births reported in Supplementary Figure2 A-C (n=169,011). This is because several included studies (Study #1, 12, 17, and 19) only had famine births but did not have control of pre- and post-famine births combined. These studies together have 6,709 famine births.

**Supplementary Figure 4B. Effect estimates of famine exposure on T2D comparing famine births with pre- and post-famine births combined after stratification by mean age at the survey**

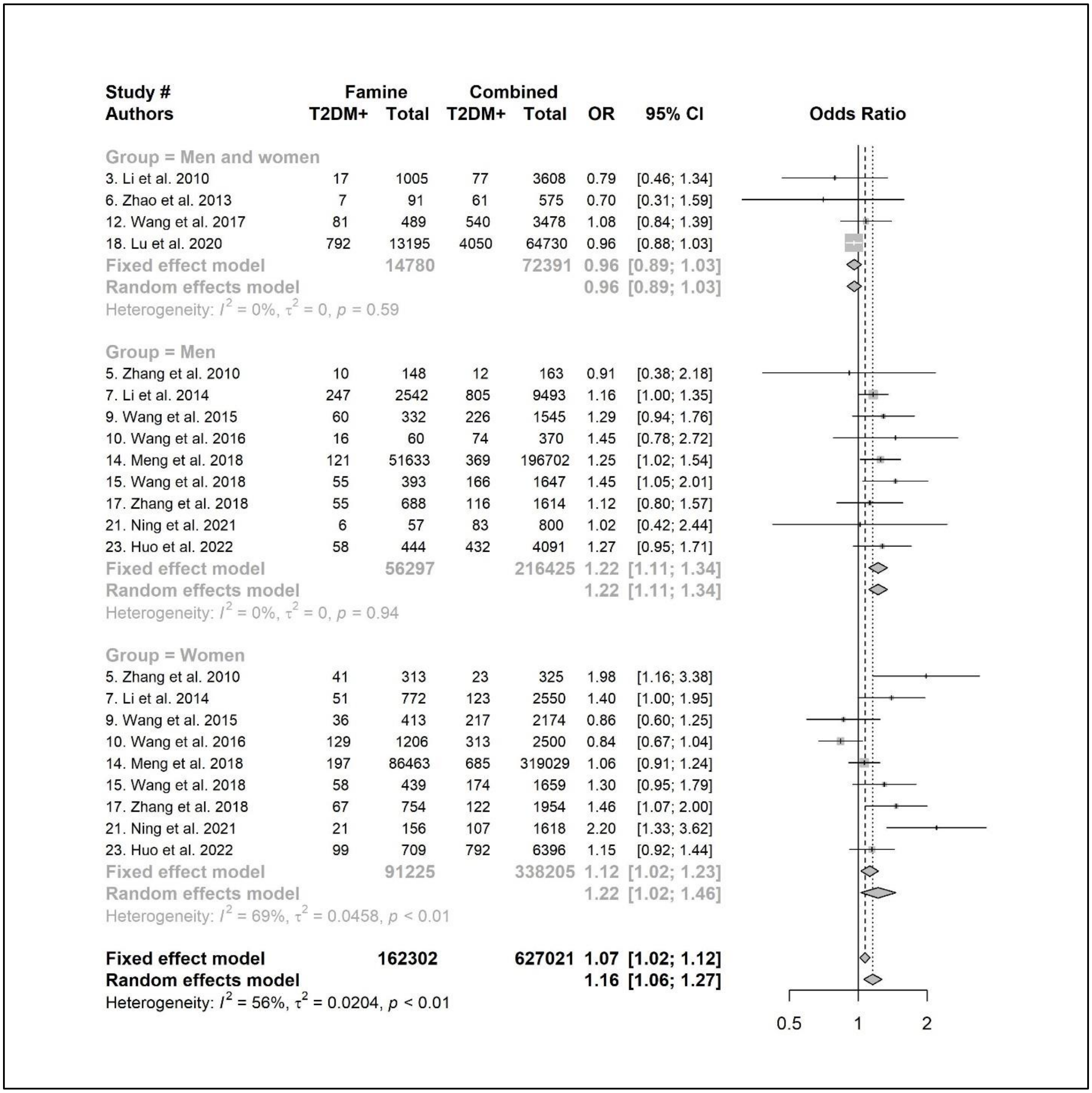

**Supplementary Figure 4C. Effect estimates of famine exposure on T2D comparing famine births with pre- and post-famine births combined after stratification by T2D measurements**

| Study # | Famine | | Combined | | | |
|---|---|---|---|---|---|---|
| Authors | T2DM+ | Total | T2DM+ | Total | OR | 95% CI |
| Group = WHO | | | | | | |
| 3. Li et al. 2010 | 17 | 1005 | 77 | 3608 | 0.79 | [0.46; 1.34] |
| 6. Zhao et al. 2013 | 7 | 91 | 61 | 575 | 0.70 | [0.31; 1.59] |
| 7. Li et al. 2014 | 298 | 3314 | 928 | 12043 | 1.18 | [1.03; 1.36] |
| 10. Wang et al. 2016 | 145 | 1266 | 387 | 2870 | 0.83 | [0.68; 1.02] |
| 15. Zhang et al. 2018 | 122 | 1442 | 238 | 3568 | 1.29 | [1.03; 1.62] |
| 21. Ning et al. 2021 | 27 | 213 | 190 | 2418 | 1.70 | [1.11; 2.62] |
| 23. Huo et al. 2022 | 157 | 1153 | 1224 | 10487 | 1.19 | [1.00; 1.43] |
| Fixed effect model | | 8484 | | 35569 | 1.12 | [1.03; 1.22] |
| Random effects model | | | | | 1.11 | [0.94; 1.32] |
| Heterogeneity: $I^2 = 66\%$, $\tau^2 = 0.0290$, $p < 0.01$ | | | | | | |
| Group = ADA | | | | | | |
| 5. Zhang et al. 2010 | 51 | 461 | 35 | 488 | 1.61 | [1.03; 2.53] |
| 9. Wang et al. 2015 | 96 | 745 | 443 | 3719 | 1.09 | [0.86; 1.39] |
| 11. Wang et al. 2017 | 81 | 489 | 540 | 3478 | 1.08 | [0.84; 1.39] |
| 14. Wang et al. 2018 | 113 | 832 | 340 | 3306 | 1.37 | [1.09; 1.72] |
| 18. Lu et al. 2020 | 792 | 13195 | 4050 | 64730 | 0.96 | [0.88; 1.03] |
| Fixed effect model | | 15722 | | 75721 | 1.02 | [0.95; 1.09] |
| Random effects model | | | | | 1.14 | [0.96; 1.37] |
| Heterogeneity: $I^2 = 70\%$, $\tau^2 = 0.0259$, $p < 0.01$ | | | | | | |
| Group = ICD-10 | | | | | | |
| 13. Meng et al. 2018 | 318 | 138096 | 1054 | 515731 | 1.13 | [0.99; 1.28] |
| Fixed effect model | | 138096 | | 515731 | 1.13 | [0.99; 1.28] |
| Random effects model | | | | | 1.13 | [0.99; 1.28] |
| Heterogeneity: not applicable | | | | | | |
| **Fixed effect model** | | **162302** | | **627021** | **1.06** | **[1.01; 1.12]** |
| **Random effects model** | | | | | **1.12** | **[1.02; 1.24]** |
| Heterogeneity: $I^2 = 66\%$, $\tau^2 = 0.0179$, $p < 0.01$ | | | | | | |

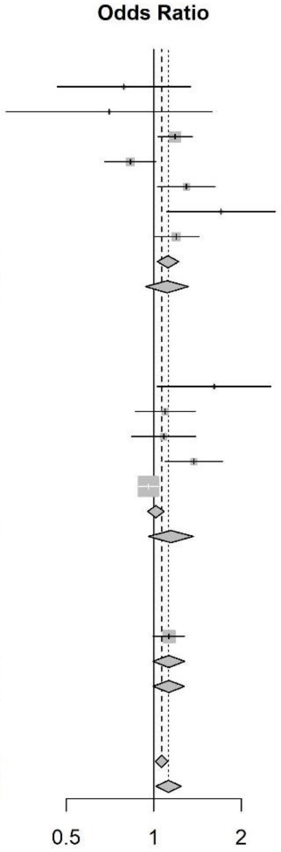

**Supplementary Figure 4D. Effect estimates of famine exposure on T2D comparing famine births with pre- and post-famine births after stratification by famine intensity**

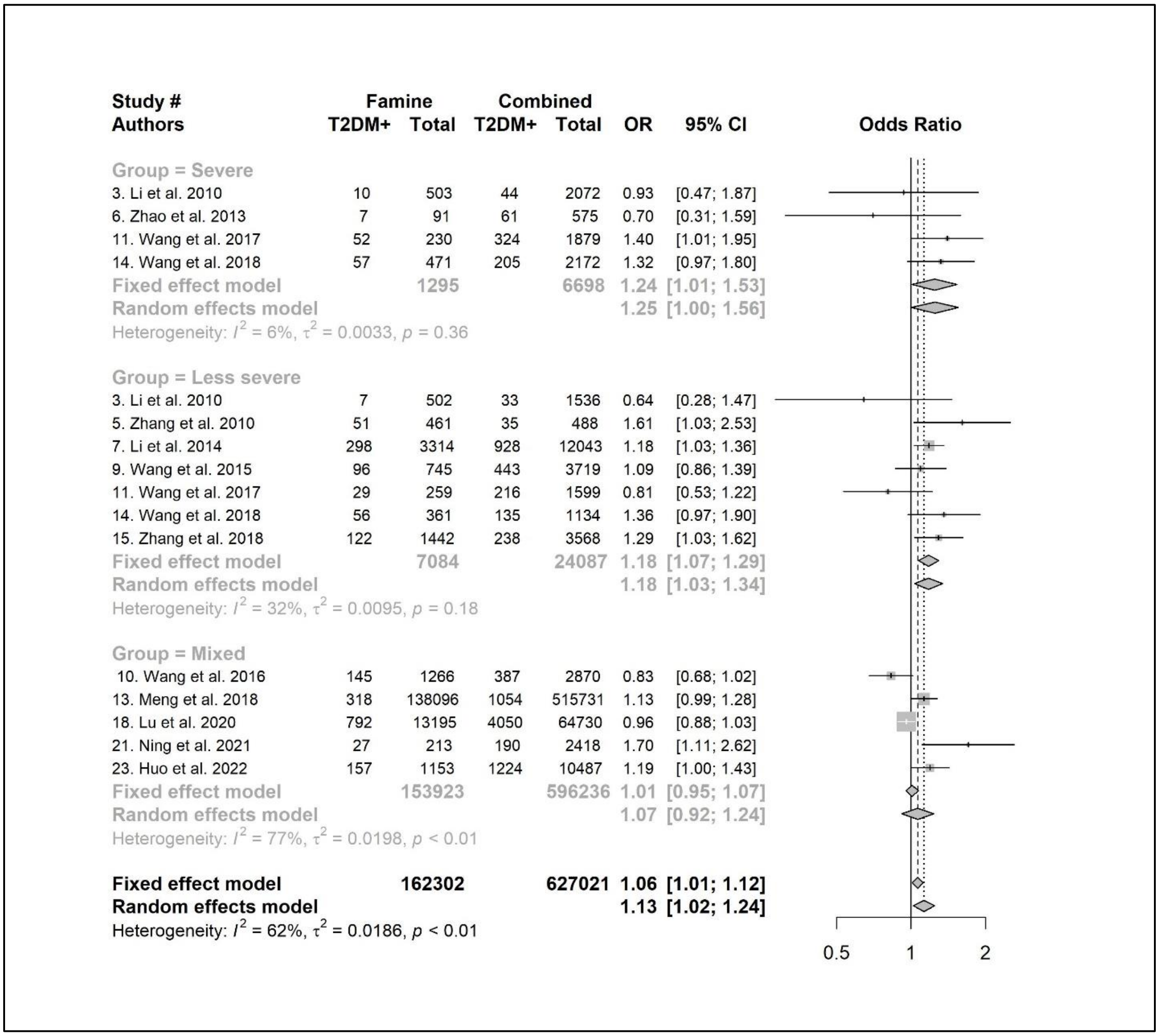

**Supplementary Figure 4E. Effect estimates of famine exposure on T2D comparing famine births with pre- and post-famine births combined after stratification by residence**

| Study # Authors | Famine T2DM+ | Famine Total | Combined T2DM+ | Combined Total | OR | 95% CI |
|---|---|---|---|---|---|---|
| **Group = Mixed** | | | | | | |
| 3. Li et al. 2010 | 17 | 1005 | 77 | 3608 | 0.79 | [0.46; 1.34] |
| 9. Wang et al. 2015 | 96 | 745 | 443 | 3719 | 1.09 | [0.86; 1.39] |
| 12. Wang et al. 2017 | 81 | 489 | 540 | 3478 | 1.08 | [0.84; 1.39] |
| 14. Meng et al. 2018 | 318 | 138096 | 1054 | 515731 | 1.13 | [0.99; 1.28] |
| 15. Wang et al. 2018 | 113 | 832 | 340 | 3306 | 1.37 | [1.09; 1.72] |
| 17. Zhang et al. 2018 | 122 | 1442 | 238 | 3568 | 1.29 | [1.03; 1.62] |
| 18. Lu et al. 2020 | 792 | 13195 | 4050 | 64730 | 0.96 | [0.88; 1.03] |
| 21. Ning et al. 2021 | 27 | 213 | 190 | 2418 | 1.70 | [1.11; 2.62] |
| **Fixed effect model** | | **156017** | | **600558** | **1.05** | **[0.99; 1.11]** |
| **Random effects model** | | | | | **1.14** | **[1.00; 1.29]** |
| Heterogeneity: $I^2 = 67\%$, $\tau^2 = 0.0182$, $p < 0.01$ | | | | | | |
| **Group = Urban** | | | | | | |
| 5. Zhang et al. 2010 | 51 | 461 | 35 | 488 | 1.61 | [1.03; 2.53] |
| 6. Zhao et al. 2013 | 7 | 91 | 61 | 575 | 0.70 | [0.31; 1.59] |
| 7. Li et al. 2014 | 298 | 3314 | 928 | 12043 | 1.18 | [1.03; 1.36] |
| 10. Wang et al. 2016 | 145 | 1266 | 387 | 2870 | 0.83 | [0.68; 1.02] |
| **Fixed effect model** | | **5132** | | **15976** | **1.07** | **[0.96; 1.20]** |
| **Random effects model** | | | | | **1.06** | **[0.79; 1.43]** |
| Heterogeneity: $I^2 = 76\%$, $\tau^2 = 0.0563$, $p < 0.01$ | | | | | | |
| **Group = Rural** | | | | | | |
| 23. Huo et al. 2022 | 157 | 1153 | 1224 | 10487 | 1.19 | [1.00; 1.43] |
| **Fixed effect model** | | **1153** | | **10487** | **1.19** | **[1.00; 1.43]** |
| **Random effects model** | | | | | **1.19** | **[1.00; 1.43]** |
| Heterogeneity: not applicable | | | | | | |
| **Fixed effect model** | | **162302** | | **627021** | **1.06** | **[1.01; 1.12]** |
| **Random effects model** | | | | | **1.12** | **[1.02; 1.24]** |
| Heterogeneity: $I^2 = 66\%$, $\tau^2 = 0.0179$, $p < 0.01$ | | | | | | |

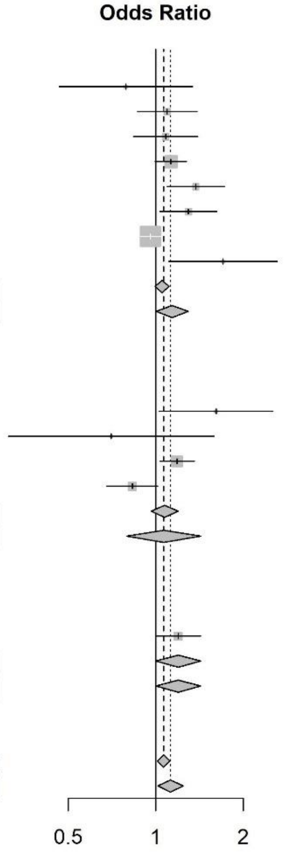

**Supplementary Figure 4F. Effect estimates of famine exposure on T2D comparing famine births with pre- and post-famine births combined stratified by publication language**

| Study # Authors | Famine T2DM+ | Famine Total | Combined T2DM+ | Combined Total | OR | 95% CI |
|---|---|---|---|---|---|---|
| **Group = English** | | | | | | |
| 3. Li et al. 2010 | 17 | 1005 | 77 | 3608 | 0.79 | [0.46; 1.34] |
| 9. Wang et al. 2015 | 96 | 745 | 443 | 3719 | 1.09 | [0.86; 1.39] |
| 10. Wang et al. 2016 | 145 | 1266 | 387 | 2870 | 0.83 | [0.68; 1.02] |
| 12. Wang et al. 2017 | 81 | 489 | 540 | 3478 | 1.08 | [0.84; 1.39] |
| 14. Meng et al. 2018 | 318 | 138096 | 1054 | 515731 | 1.13 | [0.99; 1.28] |
| 15. Wang et al. 2018 | 113 | 832 | 340 | 3306 | 1.37 | [1.09; 1.72] |
| 17. Zhang et al. 2018 | 122 | 1442 | 238 | 3568 | 1.29 | [1.03; 1.62] |
| 18. Lu et al. 2020 | 792 | 13195 | 4050 | 64730 | 0.96 | [0.88; 1.03] |
| 21. Ning et al. 2021 | 27 | 213 | 190 | 2418 | 1.70 | [1.11; 2.62] |
| 23. Huo et al. 2022 | 157 | 1153 | 1224 | 10487 | 1.19 | [1.00; 1.43] |
| **Fixed effect model** | | **158436** | | **613915** | **1.04** | **[0.99; 1.10]** |
| **Random effects model** | | | | | **1.11** | **[0.99; 1.24]** |
| Heterogeneity: $I^2 = 68\%$, $\tau^2 = 0.0187$, $p < 0.01$ | | | | | | |
| **Group = Chinese** | | | | | | |
| 5. Zhang et al. 2010 | 51 | 461 | 35 | 488 | 1.61 | [1.03; 2.53] |
| 6. Zhao et al. 2013 | 7 | 91 | 61 | 575 | 0.70 | [0.31; 1.59] |
| 7. Li et al. 2014 | 298 | 3314 | 928 | 12043 | 1.18 | [1.03; 1.36] |
| **Fixed effect model** | | **3866** | | **13106** | **1.20** | **[1.05; 1.36]** |
| **Random effects model** | | | | | **1.21** | **[0.91; 1.62]** |
| Heterogeneity: $I^2 = 40\%$, $\tau^2 = 0.0296$, $p = 0.19$ | | | | | | |
| **Fixed effect model** | | **162302** | | **627021** | **1.06** | **[1.01; 1.12]** |
| **Random effects model** | | | | | **1.12** | **[1.02; 1.24]** |
| Heterogeneity: $I^2 = 66\%$, $\tau^2 = 0.0179$, $p < 0.01$ | | | | | | |

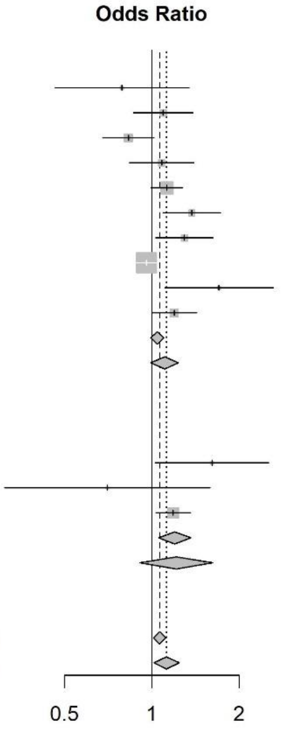

**Supplementary Figure 5. Funnel plot of effect estimates of famine exposure on T2D comparing famine births with pre- and post-famine births combined**

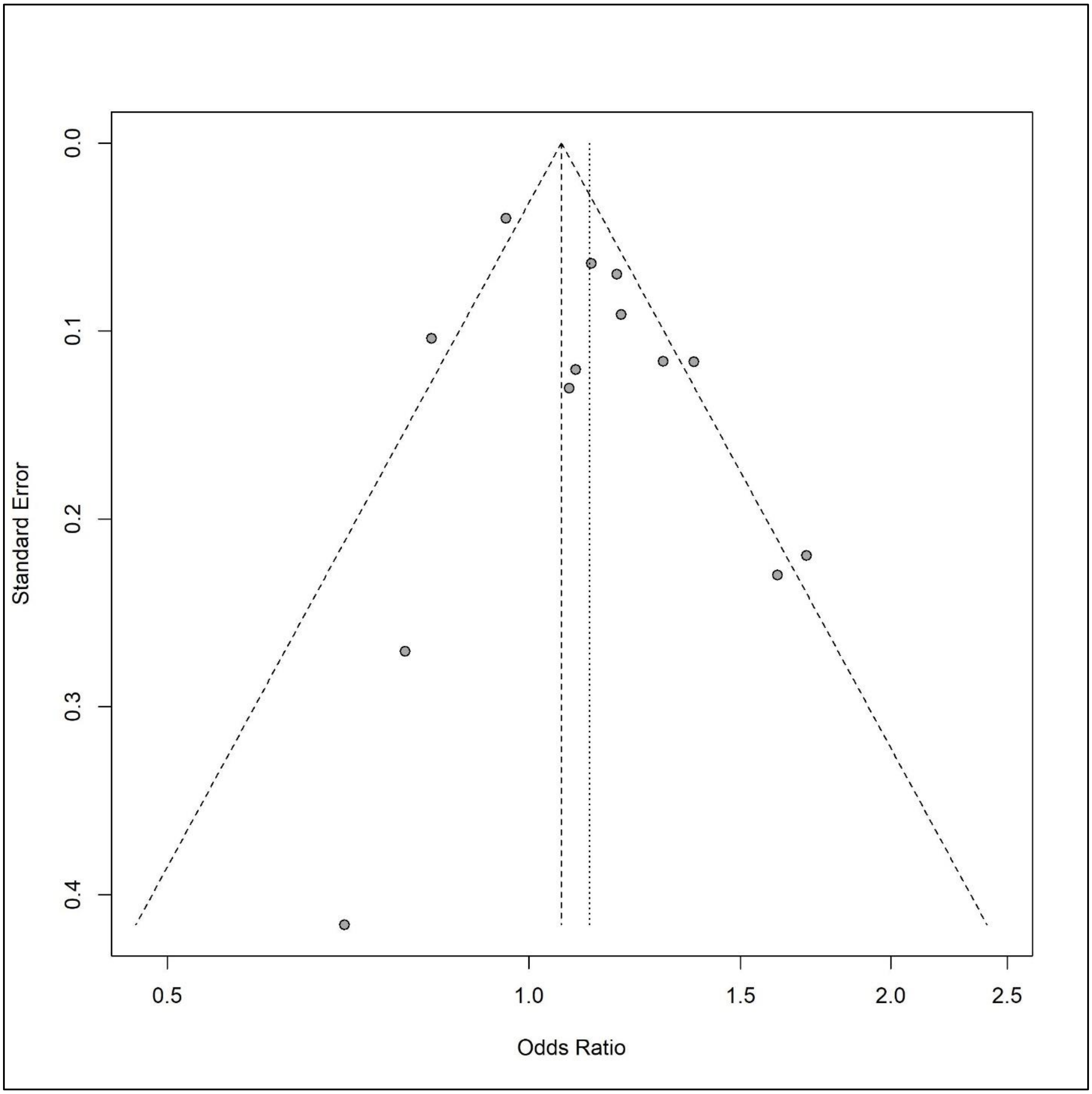

**Supplementary Reference List. Chinese famine studies on health outcomes**

The search was conducted in English databases (PubMed, Embase, and Web of Science) and Chinese databases (Chinese Wanfang Data, Chinese National Knowledge Infrastructure) on December 10th, 2021. They were listed by year of publication.

2021 (Total: 32)

Tao, Boni, et al. "Fetal exposure to the Great Chinese Famine and risk of ischemic stroke in midlife." European Journal of Neurology 28.4 (2021): 1244-1252.

VanEvery, Hannah, et al. "In utero and early life exposure to the Great Chinese Famine and risk of rheumatoid arthritis in adulthood." Arthritis & Rheumatology 73.4 (2021): 596-603.

Liu, Lin, et al. "Early-life exposure to the Chinese famine and risk of carotid intima-media thickness increased in adulthood." Nutrition, Metabolism and Cardiovascular Diseases 31.3 (2021): 841-848.

Wang, Bin, et al. "Early-life exposure to the Chinese famine, genetic susceptibility and the risk of type 2 diabetes in adulthood." Diabetologia (2021): 1-9.

Zhang, Lin, et al. "Individual and combined association analysis of famine exposure and serum uric acid with hypertension in the mid-aged and older adult: a population-based cross-sectional study." BMC Cardiovascular Disorders 21.1 (2021): 1-14.

Guo, Jiahui, et al. "Exposure to the Chinese Great Famine in early life and thyroid function and disorders in adulthood: a cross-sectional study." Thyroid 31.4 (2021): 563-571.

Rong, Hongguo, et al. "Exposure to Chinese famine in early life and the risk of sensory impairment in adulthood." J Epidemiol Community Health 75.1 (2021): 16-21.

Li, Shuxia, et al. "Differential Regulation of the DNA Methylome in Adults Born During the Great Chinese Famine in 1959-1961." Available at SSRN 3763755.

Li, Jinhu, and Nidhiya Menon. "Echo Effects of Health Shocks: The Intergenerational Consequences of Prenatal and Early-Life Malnutrition during the Great Leap Forward Famine in China." The Journal of Development Studies (2021): 1-28.

Li, Yaru, and Zhongxin Hong. "Exposure to the Chinese famine in early life and self-reported arthritis risk in adulthood." Psychology, Health & Medicine (2021): 1-10.

Li, Yanan, and Naveen Sunder. "What Doesn't Kill Her, Will Make Her Depressed." Economics & Human Biology (2021): 101064.

Li, Wei, et al. "Exposure to famine in early life and self-rated health status among Chinese adults: a cross-sectional study from the Chinese Health and Retirement Longitudinal Study (CHARLS)." BMJ open 11.10 (2021): e048214.

Li, Jie, et al. "Famine and Trajectories of Body Mass Index, Waist Circumference, and Blood Pressure in Two Generations: Results From the CHNS From 1993–2015." Hypertension (2021): HYPERTENSIONAHA-121.

Wang, Zhenghe, et al. "Early-Life Exposure to the Chinese Great Famine and Later Cardiovascular Diseases." International Journal of Public Health 66 (2021): 7.

Wang, Chenggang, and Huixia Wang. "The long-term effects of the Chinese great famine on mental health." Applied Economics Letters (2021): 1-7.

Zhang, Lin, et al. "Individual and Combined Effect of Famine Exposure and Obesity Parameters on Dyslipidemia in Mid-Aged and Older Adult: A Population-Based Cross-Sectional Study." (2021).

Jiang, Huiru, et al. "Exposure to the Great Famine in Early Life and the Risk of Obesity in Adulthood: A Report Based on the China Health and Nutrition Survey." Nutrients 13.4 (2021): 1285.

Hu, Xiang, et al. "Associations of early-life exposure to famine with abdominal fat accumulation are independent of family history of diabetes and physical activity." British Journal of Nutrition 125.8 (2021): 943-950.

Han, Xu, et al. "Cardiometabolic Traits Mediated the Relationship from Early Life Famine Exposure to Adulthood Nonalcoholic Fatty Liver Disease Risk." British Journal of Nutrition (2021): 1-27.

Huang, Yu-Qing, et al. "The relationship between famine exposure during early life and ascending aorta dilatation in adults." British Journal of Nutrition (2021): 1-8.

Huang, Yu-qing, et al. "The relationship between famine exposure during early life and carotid plaque in adulthood." European Journal of Clinical Nutrition 75.3 (2021): 546-554.

Huang, Y-Q., et al. "The relationship between famine exposure in early life and left atrial enlargement in adulthood." Journal of Human Nutrition and Dietetics 34.2 (2021): 356-364.

Meng, Ruogu, et al. "222 Long-term mortality after early famine exposure: findings from the China Kadoorie Biobank." International Journal of Epidemiology 50.Supplement_1 (2021): dyab168-434.

Wang, Yuying, et al. "Metabolites in the association between early-life famine exposure and type 2 diabetes in adulthood over a 5-year follow-up period." BMJ Open Diabetes Research and Care 9.1 (2021): e001935.

Deng, Zichen, and Maarten Lindeboom. Early-life Famine Exposure, Hunger Recall and Later-life Health. No. 21-054/V. Tinbergen Institute, 2021.

Qin, Qiaojing, et al. "Undernutrition when young and the risk of poor renal function in adulthood in women with diabetes in Shanghai, China." Journal of International Medical Research 49.5 (2021): 03000605211016671.

Guo, H. J., et al. "Association analysis of famine exposure during early life and risk of hypertension in adulthood." Zhonghua yu Fang yi xue za zhi [Chinese Journal of Preventive Medicine] 55.6 (2021): 732-736.

Liu, Chunyu, et al. "Early-life famine exposure and rheumatoid arthritis in Chinese adult populations: a retrospective cohort study." BMJ open 11.7 (2021): e043416.

Zhang, Xinyuan, et al. "In utero and childhood exposure to the Great Chinese Famine and risk of cancer in adulthood: the Kailuan Study." The American Journal of Clinical Nutrition (2021).

Wang, Bin, et al. "Early-life exposure to the Chinese famine, genetic susceptibility and the risk of type 2 diabetes in adulthood." Diabetologia (2021): 1-9.

Wang, Yu, et al. "Exposure to Chinese Famine in the Early Life, Adulthood Obesity Patterns, and the Incidence of Hypertension: A 22-Year Cohort Study." Annals of Nutrition and Metabolism (2021): 1-7.

Cheng, Mingwang, Zhouxiang Wang, and Ning Neil Yu. "Long-Term Mental Health Costs of the Great Chinese Famine." Available at SSRN 3922174 (2021).

2020 (Total: 32)

Cheng, Qu, et al. "Prenatal and early-life exposure to the Great Chinese Famine increased the risk of tuberculosis in adulthood across two generations." *Proceedings of the National Academy of Sciences* 117.44 (2020): 27549-27555.

Shi, Zumin, et al. "Early life exposure to 1959-1961 Chinese famine exacerbates association between diabetes and cardiovascular disease." *Journal of Diabetes* 12.2 (2020): 134-141.

Zhang, Hao, et al. "Early life famine exposure to the Great Chinese Famine in 1959–1961 and subsequent pregnancy loss: a population-based study." *BJOG: An International Journal of Obstetrics & Gynaecology* 127.1 (2020): 39-45.

Zhang, Wenqiang, and Rongsheng Luan. "Early-life exposure to the Chinese famine of 1959–61 and risk of Hyperuricemia: results from the China health and retirement longitudinal study." *BMC public health* 20.1 (2020): 15.

Yan, Shiwei, et al. "Prenatal exposure to the Chinese famine and the risk of metabolic syndrome in adulthood across consecutive generations." *European Journal of Clinical Nutrition* (2020): 1-8.

Guo, Jiahui, et al. "Exposure to the Chinese Great Famine in early life and thyroid function and disorders in adulthood: a cross-sectional study." *Thyroid* ja (2020).

Song, Chao, et al. "Fetal Exposure to Chinese Famine Increases Obesity Risk in Adulthood." *International Journal of Environmental Research and Public Health* 17.10 (2020): 3649.

Zhang, Yan, et al. "Exposure to Chinese Famine in Fetal Life and the Risk of Dysglycemiain Adulthood." *International Journal of Environmental Research and Public Health* 17.7 (2020): 2210.

Lv, Shiqi, et al. "Association between exposure to the Chinese famine during early life and the risk of chronic kidney disease in adulthood." *Environmental Research* (2020): 109312.

Meng, Ruogu, et al. "Early famine exposure and adult disease risk based on a 10-year prospective study of Chinese adults." *Heart* 106.3 (2020): 213-220.

Li, Yun, et al. "In utero exposure to the Great Chinese Famine and risk of intracerebral hemorrhage in midlife." *Neurology* 94.19 (2020): e1996-e2004.

Peng, Yue, et al. "Association of Exposure to Chinese Famine in Early Life with the Risk of Metabolic Syndrome in Adulthood." *Annals of Nutrition and Metabolism* 76.2 (2020): 140-146.

Na, HUANG Li, et al. "Association between Chinese Famine Exposure and the Risk of Overweight/Obesity and Abdominal Obesity in Laterlife: A Cross-sectional Study." *Biomedical and Environmental Sciences* 33.2 (2020): 133-137.

Fang, Zhe, et al. "Association Between Fetal Exposure to Famine and Anthropometric Measures in Adulthood: A Regression Discontinuity Approach." *Obesity* 28.5 (2020): 962-969.

Wang, Zhenghe, et al. "DNA methylation of the INSR gene as a mediator of the association between prenatal exposure to famine and adulthood waist circumference." *Scientific reports* 10.1 (2020): 1-8.

He, Shulan, et al. "Early-life exposure to famine and late-life depression: Does leukocyte telomere length mediate the association?." *Journal of Affective Disorders* (2020).

Huang, Y-Q., et al. "The relationship between famine exposure in early life and left atrial enlargement in adulthood." *Journal of Human Nutrition and Dietetics* (2020).

Du, Rui, et al. "Early-Life Famine Exposure and Risk of Cardiovascular Diseases in Later Life: Findings From the REACTION Study." *Journal of the American Heart Association* 9.7 (2020): e014175.

Wang, Yuying, et al. "Association between famine exposure in early life with insulin resistance and beta cell dysfunction in adulthood." *Nutrition & Diabetes* 10.1 (2020): 1-8.

Qi, Hongyan, et al. "Early life famine exposure, adulthood obesity patterns and the risk of nonalcoholic fatty liver disease." *Liver International* 40.11 (2020): 2694-2705.

Lu, Jieli, et al. "Early Life Famine Exposure, Ideal Cardiovascular Health Metrics, and Risk of Incident Diabetes: Findings From the 4C Study." *Diabetes Care* (2020).

Hu X, Wen J, Yu W, Yang L, Pan W, Xu K, Chen X, Li Q, Chen G, Gu X. Associations of early-life exposure to famine with abdominal fat accumulation are independent of family history of diabetes and physical activity. Br J Nutr. 2020 Sep 2:1-8. doi: 10.1017/S0007114520003414. Epub ahead of print. PMID: 32873353.

Ding XY, Yang ZY, Zhao LY, Zhao WH. Are Lipid Profiles in Middle Age Associated with Famine Exposure during Prenatal and Early Postnatal Period? Nutrients. 2020 Jul 29;12(8):2266. doi: 10.3390/nu12082266. PMID: 32751112; PMCID: PMC7469046.

Suo, Yue, et al. "Concurrency of Early-Age Exposure to Chinese Famine and Diabetes Increases Recurrence of Ischemic Stroke." Frontiers in neurology 11 (2020).

Song, Chao, et al. "Ten SNPs May Affect Type 2 Diabetes Risk in Interaction with Prenatal Exposure to Chinese Famine." Nutrients 12.12 (2020): 3880.

Jiang, Wenbo, et al. "Prenatal famine exposure and estimated glomerular filtration rate across consecutive generations: association and epigenetic mediation in a population-based cohort study in Suihua China." Aging (Albany NY) 12.12 (2020): 12206.

Sun, Yaoyao, et al. "Association of MAD1L1 polymorphism (rs871925) with prenatal famine exposure and schizophrenia in a Chinese population: A case–control study." IUBMB life 72.2 (2020): 259-265.

Cui, Hanxiao, James P. Smith, and Yaohui Zhao. "Early-life deprivation and health outcomes in adulthood: Evidence from childhood hunger episodes of middle-aged and elderly Chinese." Journal of development economics 143 (2020): 102417.

Wang, Yuying, et al. "Economic status moderates the association between early-life famine exposure and hyperuricemia in adulthood." The Journal of Clinical Endocrinology & Metabolism 105.11 (2020): e3862-e3873.

QI, Hongyan, et al. "Relationship between famine exposure in early life and type 2 diabetes mellitus in adulthood." Chinese Journal of Endocrinology and Metabolism (2020): 905-911.

LIU, Xuemei, et al. "Effects of famine exposure in early life on bone mineral density measured by ultrasound in postmenopausal women." Chinese Journal of Endocrinology and Metabolism (2020): 920-925.

游玥玥, et al. "生命早期饥荒暴露与成年期高血压患病风险的关联分析." 中华流行病学杂志 41.1 (2020): 74-78.

2019 (Total: 20)

Liu, Dan, et al. "Exposure to famine during early life and abdominal obesity in adulthood: Findings from the great chinese famine during 1959–1961." *Nutrients* 11.4 (2019): 903.

Zhang, Yuxia, et al. "Exposure to Chinese famine in early life modifies the association between hyperglycaemia and cardiovascular disease." *Nutrition, Metabolism and Cardiovascular Diseases* 29.11 (2019): 1230-1236.

Wang, Zhenghe, et al. "Chinese famine exposure in infancy and metabolic syndrome in adulthood: results from the China health and retirement longitudinal study." *European journal of clinical nutrition* 73.5 (2019): 724.

Zhao, Rencheng, et al. "Association of exposure to Chinese famine in early life with the incidence of hypertension in adulthood: a 22-year cohort study." *Nutrition, Metabolism and Cardiovascular Diseases* 29.11 (2019): 1237-1244.

Shen, Luqi, et al. "Early-life exposure to severe famine is associated with higher methylation level in the IGF2 gene and higher total cholesterol in late adulthood: the Genomic Research of the Chinese Famine (GRECF) study." *Clinical epigenetics* 11.1 (2019): 88.

Wang, Zhenghe, et al. "Early-Life Exposure to the Chinese Famine Is Associated with Higher Methylation Level in the INSR Gene in Later Adulthood." *Scientific reports* 9.1 (2019): 1-9.

Wang, Nengying, et al. "Early life exposure to famine and reproductive aging among Chinese women." *Menopause* 26.5 (2019): 463-468.

Xin, Xueling, et al. "Exposure to Chinese famine in early life and the risk of dyslipidemia in adulthood." *European journal of nutrition* 58.1 (2019): 391-398.

Zhou, Jielin, et al. "The effect of Chinese famine exposure in early life on dietary patterns and chronic diseases of adults." *Public health nutrition* 22.4 (2019): 603-613.

Rong, Hongguo, et al. "Early-Life Exposure to the Chinese Famine and Risk of Cognitive Decline." *Journal of Clinical Medicine* 8.4 (2019): 484.

Ning, Feng, et al. "Famine exposure in early life and risk of metabolic syndrome in adulthood: comparisons of different metabolic syndrome definitions." *Journal of diabetes research* 2019 (2019).

Li, Jiante, et al. "A pilot study on clinicopathological features and intestinal microflora changes in colorectal cancer patients born over a nine-year period encompassing three years before and after the Great Chinese famine." *Cancer epidemiology* 59 (2019): 166-172.

Zheng, Xiaoya, et al. "EXPOSURE TO THE CHINESE FAMINE IN EARLY LIFE AND THE THYROID FUNCTION AND NODULES IN ADULTHOOD." *Endocrine Practice* 25.6 (2019): 598-604.

Tao, Tao, et al. "Association between early-life exposure to the Great Chinese Famine and poor physical function later in life: a cross-sectional study." *BMJ open* 9.7 (2019): e027450.

Cheng, Wenli, and Hui Shi. "Surviving the Famine Unscathed? An Analysis of the Long-Term Health Effects of the Great Chinese Famine." *Southern Economic Journal* 86.2 (2019): 746-772.

Chen, Chi, et al. "Famine exposure in early life is associated with visceral adipose dysfunction in adult females." *European journal of nutrition* 58.4 (2019): 1625-1633.

Yao, Huihui, and Li Li. "Famine exposure during the fetal period increased the risk of dyslipidemia in female adults." *Lipids* 54.5 (2019): 301-309.

Zong, Liyao, et al. "Exposure to famine in early life and the risk of osteoporosis in adulthood: A prospective study." *Endocrine Practice* 25.4 (2019): 299-305.

林淑贞, 周泳宏. 饥荒,性格形成与心理健康[J]. 劳动经济研究, 2019, v.7;No.37(06):38-64.

Liu, Y., et al., Effect of famine exposure during fetal period on occurence of diabetes after adult in results of Zhuang nationality. Guangxi Med J, 2019. 41(2): p. 221-224.

2018 (Total: 17)

Shi, Zumin, et al. "Early life exposure to Chinese famine modifies the association between hypertension and cardiovascular disease." *Journal of hypertension* 36.1 (2018): 54-60.

He, Ping, et al. "Prenatal malnutrition and adult cognitive impairment: a natural experiment from the 1959–1961 Chinese famine." *British Journal of Nutrition* 120.2 (2018): 198-203.

Yu, Caizheng, et al. "Victims of Chinese famine in early life have increased risk of metabolic syndrome in adulthood." *Nutrition* 53 (2018): 20-25.

Rong, Hongguo, et al. "The correlation between early stages of life exposed to Chinese famine and cognitive decline in adulthood: nutrition of adulthood plays an important role in the link?." *Frontiers in aging neuroscience* 9 (2018): 444.

Li, Yaru, et al. "Exposure to the Chinese famine in early life and depression in adulthood." *Psychology, health & medicine* 23.8 (2018): 952-957.

Zheng, Xiaoya, et al. "The great chinese famine exposure in early life and the risk of nonalcoholic fatty liver disease in adult women." *Annals of hepatology* 16.6 (2018): 901-908.

Meng, Ruogu, et al. "Prenatal famine exposure, adulthood obesity patterns and risk of type 2 diabetes." *International journal of epidemiology* 47.2 (2018): 399-408.

Zhang, Yangyu, et al. "Risk of hyperglycemia and diabetes after early-life famine exposure: A cross-sectional survey in northeastern china." *International journal of environmental research and public health* 15.6 (2018): 1125.

Xu, Hongwei, et al. "Early life exposure to China's 1959–61 famine and midlife cognition." *International journal of epidemiology* 47.1 (2018): 109-120.

Boks, M. P., et al. "Genetic vulnerability to DUSP22 promoter hypermethylation is involved in the relation between in utero famine exposure and schizophrenia." *npj Schizophrenia* 4.1 (2018): 1-8.

Wang, Zhenghe, et al. "The association between fetal-stage exposure to the China famine and risk of diabetes mellitus in adulthood: results from the China health and retirement longitudinal study." *BMC public health* 18.1 (2018): 1205.

Li, Changwei, et al. "Early-life exposure to severe famine and subsequent risk of depressive symptoms in late adulthood: the China Health and Retirement Longitudinal Study." *The British Journal of Psychiatry* 213.4 (2018): 579-586.

Chang, X., et al. "The Risks of Overweight, Obesity and Abdominal Obesity in Middle Age after Exposure to Famine in Early Life: Evidence from the China's 1959–1961 Famine." *The journal of nutrition, health & aging* 22.10 (2018): 1198-1204.

Wang, Zhenghe, et al. "Association between the Great China Famine exposure in early life and risk of arthritis in adulthood." *J Epidemiol Community Health* 72.9 (2018): 790-795.

Wang, Ningjian, et al. "Exposure to famine in early life and chronic kidney diseases in adulthood." *Nutrition & diabetes* 8.1 (2018): 1-7.

汪蒙, 宋超, 宫伟彦,等. 20 世纪 60 年代初期中国出生人群糖尿病相关基因多态性[J]. 卫生研究, 2018, 047(003):358-366.

汪蒙. 胚胎期饥荒、基因与成年糖代谢关系的研究[D].

2017 (Total: 19)

Liu, L., et al. "Exposure to famine in early life and the risk of obesity in adulthood in Qingdao: Evidence from the 1959–1961 Chinese famine." *Nutrition, Metabolism and Cardiovascular Diseases* 27.2 (2017): 154-160.

Yu, Caizheng, et al. "Exposure to the Chinese famine in early life and hypertension prevalence risk in adults." *Journal of hypertension* 35.1 (2017): 63-68.

Wu, Lei, et al. "Prenatal exposure to the Great Chinese Famine and mid-age hypertension." *PLoS One* 12.5 (2017): e0176413.

He, Dandan, et al. "Incidence of breast cancer in Chinese women exposed to the 1959–1961 great Chinese famine." *BMC cancer* 17.1 (2017): 824.

Wang, Zhenghe, et al. "Fetal and infant exposure to severe Chinese famine increases the risk of adult dyslipidemia: Results from the China health and retirement longitudinal study." *BMC Public Health* 17.1 (2017): 488.

Wang, Ningjian, et al. "The famine exposure in early life and metabolic syndrome in adulthood." *Clinical Nutrition* 36.1 (2017): 253-259.

Wang, Zhenghe, et al. "Association between exposure to the Chinese famine during infancy and the risk of self-reported chronic lung diseases in adulthood: a cross-sectional study." *BMJ open* 7.5 (2017): e015476.

Wang, Cuntong, and Yudong Zhang. "Schizophrenia in mid-adulthood after prenatal exposure to the Chinese Famine of 1959–1961." *Schizophrenia Research* 184 (2017): 21-25.

Li, Jie, et al. "Prenatal exposure to famine and the development of hyperglycemia and type 2 diabetes in adulthood across consecutive generations: a population-based cohort study of families in Suihua, China." *The American Journal of Clinical Nutrition* 105.1 (2017): 221-227.

Zhang, Zhuoni, Shige Song, and Xiaogang Wu. "Exodus from hunger: The long-term health consequences of the 1959–1961 Chinese famine." *Biodemography and social biology* 63.2 (2017): 148-166.

Liu, Lingli, et al. "Increase in the prevalence of hypertension among adults exposed to the Great Chinese Famine during early life." *Environmental health and preventive medicine* 22.1 (2017): 1-7.

Xu, Xianglong, et al. "Increase in the prevalence of arthritis in adulthood among adults exposed to Chinese famine of 1959 to 1961 during childhood: A cross-sectional survey." *Medicine* 96.13 (2017).

Xie, Shao-Hua, and Jesper Lagergren. "A possible link between famine exposure in early life and future risk of gastrointestinal cancers: Implications from age-period-cohort analysis." *International journal of cancer* 140.3 (2017): 636-645.

Wang, Ningjian, et al. "Exposure to severe famine in the prenatal or postnatal period and the development of diabetes in adulthood: an observational study." *Diabetologia* 60.2 (2017): 262-269.

Mei, Zhendong, and Jiucun Wang. "Exposure to the Chinese famine in early life and the risk of hyperuricemia in adulthood among different gender." 2017 中国长三角遗传学大会会议手册. 2017.

荣红国, and 肖荣. "Different Early Life Stages Exposed to The Chinese Famine Associated with Cognitive Decline in Adulthood: Nutrition Environment Play an Important Role in The Link?." 2017 中国营养医学发展论坛暨全军营养医学大会论文汇编 (2017).

李婷. 中国老年人生理年龄的测量[J]. 人口研究, 2017, 41(6):3-15.

王瑶, 王永红, 陈霞,等. 生命早期暴露于中国饥荒年(1959 年至 1961 年)人群认知状态的研究[J]. 重庆医科大学学报, 2015(1):41-45.

王能颖. 胎儿暴露于饥荒与卵巢早衰的关系[D]. 2017.

2016 (Total: 10)

Wang, Jing, et al. "Exposure to the Chinese famine in childhood increases type 2 diabetes risk in adults." *The Journal of nutrition* 146.11 (2016): 2289-2295.

Wang, Chao, et al. "Association between exposure to the Chinese famine in different stages of early life and decline in cognitive functioning in adulthood." *Frontiers in Behavioral Neuroscience* 10 (2016): 146.

Wang, Zhenghe, et al. "Infant exposure to Chinese famine increased the risk of hypertension in adulthood: results from the China Health and Retirement Longitudinal Study." *BMC Public Health* 16.1 (2016): 435.

Chen, Jiang-Peng, et al. "Fetal and infant exposure to the Chinese famine increases the risk of fatty liver disease in Chongqing, China." *Journal of gastroenterology and hepatology* 31.1 (2016): 200-205.

Wang, Ningjian, et al. "Exposure to famine in early life and nonalcoholic fatty liver disease in adulthood." *The Journal of Clinical Endocrinology & Metabolism* 101.5 (2016): 2218-2225.

Alimujiang, Aliya, et al. "The association between China’s Great famine and risk of breast cancer according to hormone receptor status: a hospital-based study." *Breast cancer research and treatment* 160.2 (2016): 361-369.

Xu, Hongwei, et al. "Is natural experiment a cure? Re-examining the long-term health effects of China's 1959–1961 famine." *Social Science & Medicine* 148 (2016): 110-122.

Shao-Hua, Xie, and Lagergren Jesper. "A possible link between famine exposure in early life and future risk of gastrointestinal cancers: implications from age-period-cohort analysis." (2016).

Kim S, Fleisher BM, Sun JY. The long-term health effects of fetalmalnutrition: evidence from the 1959–1961 China Great Leap Forward Famine. Health Econ 2016, Aug 19. doi: 10.1002/hec.3397

魏世超, and 陈刚. "Association between sleep quality in adulthood and exposure to Chinese Great Famine in early life: A retrospective study in he population." *中国睡眠研究会第九届学术年会汇编* (2016).

2015 (Total: 7)

Fan, Wen, and Yue Qian. "Long-term health and socioeconomic consequences of early-life exposure to the 1959–1961 Chinese Famine." *Social science research* 49 (2015): 53-69.

Tan, Chih Ming, Tan Zhibo, and Xiaobo Zhang. "Sins of the Fathers: The Intergenerational Legacy of the 1959-61 Great Chinese Famine on Children's Cognitive Development." *Available at SSRN 2409452* (2015).

Li, Jie, et al. "Multigenerational effects of parental prenatal exposure to famine on adult offspring cognitive function." *Scientific reports* 5.1 (2015): 1-8.

Wang, Ningjian, et al. "Is exposure to famine in childhood and economic development in adulthood associated with diabetes?." *The Journal of Clinical Endocrinology & Metabolism* 100.12 (2015): 4514-4523.

Li, Qiang, and Lian An. "Intergenerational health consequences of the 1959–1961 Great Famine on children in rural China." *Economics & Human Biology* 18 (2015): 27-40.

王玮. 青岛地区饥荒年出生人群高血压相关调查分析[D]. 2015.

Li Y, Zhu L, Wang J et al. The effects of exposure to the famine during early life with elevated resting heart rate in the adult. Chin J Prev Med 2015;49:600–04.

2014 (Total: 6)

Chen, Henian, Wendy N. Nembhard, and Heather G. Stockwell. "Sex-specific effects of fetal exposure to the 1959–1961 Chinese famine on risk of adult hypertension." *Maternal and child health journal* 18.3 (2014): 527-533.

Huang, Cheng, et al. "Elevated levels of protein in urine in adulthood after exposure to the Chinese famine of 1959–61 during gestation and the early postnatal period." *International journal of epidemiology* 43.6 (2014): 1806-1814.

Li Y, Han H, Chen S, Lu Y, Zhu L, Wen W, Cui L, Wu S. [Effects related to experiences of famine during early life on diabetes mellitus and impaired fasting glucose during adulthood]. Zhonghua Liu Xing Bing Xue Za Zhi. 2014 Jul;35(7):852-5. Chinese. PMID: 25294081.

Li Y, Shen Y, Ye K, Zhang D, Zhang J, Li L, Zhao Q. [Association between early life exposure to famine and damaging the liver and kidney function]. Zhonghua Liu Xing Bing Xue Za Zhi. 2014 Jul;35(7):848-51. Chinese. PMID: 25294080.

Song S. Malnutrition, sex ratio, and selection: a study based on the great leap forward famine. Hum Nat. 2014 Dec;25(4):580-95. doi: 10.1007/s12110-014-9208-1. PMID: 25129431.

Zhang J, Li Y, Li L, Li X. The effect of the famine exposure in early life of the 50-year-old adult on the levels of body fat and blood glucose. J Bengbu Med School 2014;39:99–102.

2013 (Total: 5)

Song S. Identifying the intergenerational effects of the 1959-1961 Chinese Great Leap Forward Famine on infant mortality. Econ Hum Biol. 2013 Dec;11(4):474-87. doi: 10.1016/j.ehb.2013.08.001. Epub 2013 Sep 12. PMID: 24095302.

Shi Z, Zhang C, Zhou M, Zhen S, Taylor AW. Exposure to the Chinese famine in early life and the risk of anaemia in adulthood. BMC Public Health. 2013 Oct 1;13:904. doi: 10.1186/1471-2458-13-904. PMID: 24079608; PMCID: PMC3849930.

Song S. Assessing the impact of in utero exposure to famine on fecundity: evidence from the 1959-61 famine in China. Popul Stud (Camb). 2013;67(3):293-308. doi: 10.1080/00324728.2013.774045. Epub 2013 Mar 15. PMID: 23495746.

Huang C, Phillips MR, Zhang Y, Zhang J, Shi Q, Song Z, Ding Z, Pang S, Martorell R. Malnutrition in early life and adult mental health: evidence from a natural experiment. Soc Sci

Med. 2013 Nov;97:259-66. doi: 10.1016/j.socscimed.2012.09.051. Epub 2012 Dec 20. PMID: 23313495; PMCID: PMC3726543.

Zhao, Y. "Exposure to the 1959–1961 Chinese famine in early life and the risk of chronic metabolic diseases in adulthhood." *Anhui Medical University* (2013).

2012 (Total: 6)

Wang PX, Wang JJ, Lei YX, Xiao L, Luo ZC. Impact of fetal and infant exposure to the Chinese Great Famine on the risk of hypertension in adulthood. PLoS One. 2012;7(11):e49720. doi: 10.1371/journal.pone.0049720. Epub 2012 Nov 21. PMID: 23185416; PMCID: PMC3504120.

Li QD, Li H, Li FJ, Wang MS, Li ZJ, Han J, Li QH, Ma XJ, Wang da N. Nutrition deficiency increases the risk of stomach cancer mortality. BMC Cancer. 2012 Jul 28;12:315. doi: 10.1186/1471-2407-12-315. PMID: 22838407; PMCID: PMC3443031.

Song S. Does famine influence sex ratio at birth? Evidence from the 1959-1961 Great Leap Forward Famine in China. Proc Biol Sci. 2012 Jul 22;279(1739):2883-90. doi: 10.1098/rspb.2012.0320. Epub 2012 Mar 28. PMID: 22456881; PMCID: PMC3367790.

Yang J, Wang H, Zhao Y. [Analysis on the quality of life of people born in famine years (1959 - 1961) in Chongqing city and its influencing factors]. Wei Sheng Yan Jiu. 2011 Sep;40(5):611-4. Chinese. PMID: 22043713.

Zheng X, Wang Y, Ren W, Luo R, Zhang S, Zhang JH, Zeng Q. Risk of metabolic syndrome in adults exposed to the great Chinese famine during the fetal life and early childhood. Eur J Clin Nutr. 2012 Feb;66(2):231-6. doi: 10.1038/ejcn.2011.161. Epub 2011 Oct 5. PMID: 21970943.

Gørgens, Tue, Xin Meng, and Rhema Vaithianathan. "Stunting and selection effects of famine: A case study of the Great Chinese Famine." *Journal of development Economics* 97.1 (2012): 99-111.

2011 (Total: 5)

Li Y, Jaddoe VW, Qi L, He Y, Lai J, Wang J, Zhang J, Hu Y, Ding EL, Yang X, Hu FB, Ma G. Exposure to the Chinese famine in early life and the risk of hypertension in adulthood. J Hypertens. 2011 Jun;29(6):1085-92. doi: 10.1097/HJH.0b013e328345d969. PMID: 21546877.

Li Y, Jaddoe VW, Qi L, He Y, Wang D, Lai J, Zhang J, Fu P, Yang X, Hu FB. Exposure to the chinese famine in early life and the risk of metabolic syndrome in adulthood. Diabetes Care. 2011 Apr;34(4):1014-8. doi: 10.2337/dc10-2039. Epub 2011 Feb 10. PMID: 21310886; PMCID: PMC3064015.

Huang C, Li Z, Narayan KM, Williamson DF, Martorell R. Bigger babies born to women survivors of the 1959-1961 Chinese famine: a puzzle due to survival selection? J Dev Orig Health Dis. 2010 Dec;1(6):412-8. doi: 10.1017/S2040174410000504. PMID: 25142012.

Mu R, Zhang X. Why does the Great Chinese Famine affect the male and female survivors differently? Mortality selection versus son preference. Econ Hum Biol. 2011 Jan;9(1):92-105. doi: 10.1016/j.ehb.2010.07.003. Epub 2010 Aug 3. PMID: 20732838.

马光荣. "中国大饥荒对健康的长期影响: 来自 CHARLS 和县级死亡率历史数据的证据." *世界经济* 4 (2011): 104-123.

2010 (Total: 7)

Huang C, Li Z, Wang M, Martorell R. Early life exposure to the 1959-1961 Chinese famine has long-term health consequences. J Nutr. 2010 Oct;140(10):1874-8. doi: 10.3945/jn.110.121293. Epub 2010 Aug 11. PMID: 20702751.

Li Y, He Y, Qi L, Jaddoe VW, Feskens EJ, Yang X, Ma G, Hu FB. Exposure to the Chinese famine in early life and the risk of hyperglycemia and type 2 diabetes in adulthood. Diabetes. 2010 Oct;59(10):2400-6. doi: 10.2337/db10-0385. Epub 2010 Jul 9. PMID: 20622161; PMCID: PMC3279550.

Wang Y, Wang X, Kong Y, Zhang JH, Zeng Q. The Great Chinese Famine leads to shorter and overweight females in Chongqing Chinese population after 50 years. Obesity (Silver Spring). 2010 Mar;18(3):588-92. doi: 10.1038/oby.2009.296. Epub 2009 Sep 24. PMID: 19779478.

Almond, Douglas, et al. "Long-term effects of early-life development: Evidence from the 1959 to 1961 china famine." *The economic consequences of demographic change in East Asia*. University of Chicago Press, 2010. 321-345.

Song, Shige. "Mortality consequences of the 1959–1961 Great Leap Forward famine in China: Debilitation, selection, and mortality crossovers." *Social Science & Medicine* 71.3 (2010): 551-558.

Fung, Winnie, and Wei Ha. "Intergenerational effects of the 1959–61 China famine." *Risk, shocks, and human development*. Palgrave Macmillan, London, 2010. 222-254.

Li T, Liu D, Wang Y, Zhao Y, Guan Y. Hyperglycaemia survey during adulthood among people born in famine years. The Seventh National Conference of Maternal and Child Nutrition of Chinese Nutrition Society, Conference date: 11 October 2010. Nanjing, China: Chinese Nutrition Society, 2010.

2009 (Total: 6)

Song S. Does famine have a long-term effect on cohort mortality? Evidence from the 1959-1961 great leap forward famine in China. J Biosoc Sci. 2009 Jul;41(4):469-91. doi: 10.1017/S0021932009003332. Epub 2009 Mar 23. PMID: 19302727.

Song S, Wang W, Hu P. Famine, death, and madness: schizophrenia in early adulthood after prenatal exposure to the Chinese Great Leap Forward Famine. Soc Sci Med. 2009 Apr;68(7):1315-21. doi: 10.1016/j.socscimed.2009.01.027. Epub 2009 Feb 14. PMID: 19232455.

Xu MQ, Sun WS, Liu BX, Feng GY, Yu L, Yang L, He G, Sham P, Susser E, St Clair D, He L. Prenatal malnutrition and adult schizophrenia: further evidence from the 1959-1961 Chinese famine. Schizophr Bull. 2009 May;35(3):568-76. doi: 10.1093/schbul/sbn168. Epub 2009 Jan 20. PMID: 19155344; PMCID: PMC2669578.

Meng, Xin, and Nancy Qian. *The long term consequences of famine on survivors: evidence from a unique natural experiment using China's great famine*. No. w14917. National Bureau of Economic Research, 2009.

关蕴良, 王永红, 李廷玉,等. 重庆市饥荒时期出生人群代谢综合征现时患病情况调查[J]. 医学争鸣, 2009, 030(024):3173-3177.

Liu D, Wang Y, Li T, Zhao Y. Attack rate of diabetes mellitus is significantly high in adults, born in 1960 (intermediate stage of famine times). J Chongqing Med Univ 2009;34:1712–14.

2008 (Total: 1)

Yang Z, Zhao W, Zhang X, Mu R, Zhai Y, Kong L, Chen C. Impact of famine during pregnancy and infancy on health in adulthood. Obes Rev. 2008 Mar;9 Suppl 1:95-9. doi: 10.1111/j.1467-789X.2007.00447.x. PMID: 18307708.

2007 (Total: 2)

Kin CF, Shan WS, Shun LJ, Chung LP, Jean W. Experience of famine and bone health in post-menopausal women. Int J Epidemiol. 2007 Oct;36(5):1143-50. doi: 10.1093/ije/dym149. Epub 2007 Jul 26. PMID: 17660196.

Chen, Yuyu, and Li-An Zhou. "The long-term health and economic consequences of the 1959–1961 famine in China." *Journal of health economics* 26.4 (2007): 659-681.

2006 (Total: 2)

Zhao WH, Yang ZX, Zhai Y, Kong LZ, Chen CM. [Effect of nutritional status during infancy and childhood on the risk of overweight and obesity in adulthood]. Zhonghua Liu Xing Bing Xue Za Zhi. 2006 Aug;27(8):647-50. Chinese. PMID: 17172100.

Luo, Zhehui, Ren Mu, and Xiaobo Zhang. "Famine and overweight in China." *Review of Agricultural Economics* 28.3 (2006): 296-304.

2005 (Total: 2)

St Clair D, Xu M, Wang P, Yu Y, Fang Y, Zhang F, Zheng X, Gu N, Feng G, Sham P, He L. Rates of adult schizophrenia following prenatal exposure to the Chinese famine of 1959-1961. JAMA. 2005 Aug 3;294(5):557-62. doi: 10.1001/jama.294.5.557. PMID: 16077049.

Cai Y, Feng W. Famine, social disruption, and involuntary fetal loss: evidence from Chinese survey data. Demography. 2005 May;42(2):301-22. doi: 10.1353/dem.2005.0010. PMID: 15986988.

1998 (Total: 1)

Zhou L, Corruccini RS. Enamel hypoplasias related to famine stress in living Chinese. Am J Hum Biol. 1998;10(6):723-733. doi: 10.1002/(SICI)1520-6300(1998)10:6. PMID: 28561411.
